# Sex differences in the secular change in waist circumference relative to body mass index in the Americas and England from 1997 to 2020

**DOI:** 10.1101/2023.12.10.23299756

**Authors:** Luz M Sánchez-Romero, Janine Sagaceta-Mejía, Jennifer S Mindell, Álvaro Passi-Solar, Antonio Bernabé-Ortiz, Lizbeth Tolentino-Mayo, Alison Moody, Shaun Scholes

## Abstract

**Objective:** To quantify changes over time in waist circumference (WC) relative to body mass index (BMI) by sex in the Americas (U*nited* S*tates of America*, Mexico, Chile, Peru) and England.

**Methods:** Data from adults aged 25-64 years between 1997 and 2020 was analysed; US data was stratified by racial-ethnic groups. Sex-specific BMI and WC means, and obesity and abdominal obesity prevalence, were compared between the first and last surveys. Using data from all survey years, secular changes across the BMI and WC distributions were estimated applying quantile regression models. BMI was added as a predictor of WC to estimate secular changes in WC relative to BMI. Interaction terms were included in all models to evaluate differences by sex.

**Results:** BMI and WC (except Peru) showed larger secular increases at the upper-tails of the distributions in both sexes. Increases at the 50^th^ and 75^th^ WC centiles relative to BMI were more pronounced in women than in men, with larger increases in US non-Hispanic whites and in England. In men, increases in WC independently of BMI were most evident in Mexico.

**Conclusions:** Disease risk associated with visceral fat, is potentially underestimated by national surveillance efforts that quantify *secular* changes only in BMI.

## Study Importance

### What is already known about this subject?

- Body mass index (BMI) alone is not sufficient as an anthropometric indicator to assess health-related risks.
- Waist circumference (WC) and BMI are highly positively correlated at the individual level.

### What are the new findings in your manuscript?

- Mixed patterns in the Americas (US, Mexico, Chile, Peru) and in England in adults aged 25-64 years were observed for secular changes in WC relative to BMI between 1997 and 2020.
- Increases in WC relative to BMI were observed, especially, but not exclusively, in women.

### How might your results change the direction of research?

- Disease risk, associated with visceral fat, is underestimated by national surveillance efforts that quantify secular changes only in BMI.
- More investigation is needed to identify the key modifiable factors underlying recent increases in WC relative to BMI.

## INTRODUCTION

Obesity, defined as body mass index (BMI) ≥30kg/m^2^, has increased worldwide in the past few decades. More than 1.9 billion adults aged 18 years and older had overweight in 2016, and of these, over 650 million adults had obesity (1). BMI, a marker of general adiposity, has been the anthropometric indicator most commonly used to assess trends in body fatness at the population level (2). BMI is also frequently used to assess cardiometabolic risk and disease burden associated with body fatness (3–5).

BMI alone is not sufficient as an indicator to assess health-related risks properly; its limitations as a measure of body fatness have been widely recognised. BMI does not assess the distribution of body fat, and the adverse consequences of obesity may be strongly associated with the amount of visceral fat (6). Waist circumference (WC), a marker of abdominal adiposity, is acknowledged as a simple, more sensitive measure that enables better assessment of visceral adiposity associated health risk (7); WC is now recommended as a complementary measure alongside BMI in epidemiology studies and clinical practice (8).

WC and BMI are highly positively correlated at the individual level (∼0.9) (6). As such, secular increases in mean WC at the population level may have been expected to have largely occurred because of the increases in BMI during the same period. However, an emerging global concern is a secular change in the BMI-WC association, namely a recent shift to increasing abdominal obesity (indicated by higher WC) independently of BMI (9).

Only a few studies have assessed secular changes in WC relative to BMI (6, 10–12) or body weight (13), and whether such changes differed by sex, countries or racial-ethnic groups (9, 14). Freedman and Ford, using United States of America (US) data, reported that the increase in mean WC between 1999-2000 and 2011-2012 was independent of BMI increases during that same time period in women but not in men (6). Albrecht et al., using data spanning more than a decade, reported disproportionate increases in mean WC relative to BMI in the populations of the US, Mexico, China, and England, particularly in women aged 20-29 years (14). Among participants in the 1946, 1958, and 1970 British birth cohort studies, Johnson et al. observed higher increases in WC independent of BMI in mid-adulthood in women but not in men (15).

It is relevant to update and expand on these research efforts by exploring recent secular changes in WC relative to BMI in other countries to assess whether the findings described above are also observed in countries such as Chile and Peru, where the increase in obesity has been more rapid in recent years. Using data from the Americas (US, Mexico, Chile, and Peru) and England, our objective was to quantify secular changes in BMI, WC, and WC relative to BMI within each country, and explore differences by sex.

## METHODS

Data were obtained from five nationally-representative health examination surveys (HES). All countries’ surveys collect cross-sectional data from the civilian non-institutionalised population using face-to-face interviews and direct measurements of height, weight and WC. Table S1 shows a detailed description of the HES design and methods. Further information on a number of these surveys, compiled by the Encuestas de Salud de las Americas y el Reino Unido (ESARU) network of HES researchers from the Americas and the UK, is available in Mindell et al. (16).

### Chile

Chilean data was obtained from the National Health Survey “*Encuesta Nacional de Salud*” (ENS) 2003, 2010 and 2017 (17). ENS aims to provide information on prioritised health conditions and their respective treatments in the Chilean adult population. The 2003 ENS sample was stratified with a random sub-sample from participants of the Quality of Life and Health Survey with fresh samples selected for ENS 2010 and 2017. Persons aged 65 years and over were oversampled, with one eligible person sampled per household. Study protocols and ethical consent forms were approved by the ethics committee of the Pontificia Universidad Católica de Chile (PUC) and the Chilean Ministry of Health. Persons selected for inclusion provided informed and signed consent before participation.

### Mexico

For Mexico, we used the National Health and Nutrition Survey “*Encuesta Nacional de Salud y Nutrición*” (ENSANUT) 2006, 2012 and 2018. Conducted every six years, ENSANUT characterises the health and nutritional status of the Mexican population (18). ENSANUT is a cross-sectional, multi-stage, stratified, cluster sample conducted by Mexico’s Instituto Nacional de Nutrición (INSP) (National Institute of Public Health). Data collection was approved by the INSP Internal Review Board and all participants gave informed consent.

### Peru

For Peru, we included The Demographic and Family Health Survey *“Encuesta Demográfica y de Salud Familiar”* (ENDES) 2018, 2019 and 2020. ENDES provides health information to characterise communicable and non-communicable diseases in the Peruvian population (ENDES surveys before 2018 assessed only the health status of women of reproductive age and of children aged under five-years).

### United States

US data was obtained from The National Health and Nutrition Examination Survey (NHANES), conducted by the National Center for Health Statistics. NHANES uses a complex, multistage sample design. Survey participants complete in-home interviews followed by medical examinations in mobile examination centres. The National Center for Health Statistics ethics review board approved the survey, and participants gave informed consent. We used data from the ten 2-year cycles conducted continuously from 1999-2000 through 2017-2018. For this study we combined consecutive 2-year cycles into pooled 4-year time-periods to increase sample sizes.

### England

For England, data was obtained from the Health Survey for England (HSE). The HSE annually draws a new nationally-representative sample of people living in private households using multistage stratified probability sampling (19). All adults in selected households are eligible for interview. Relevant committees granted research ethics approval for the survey. Participants gave verbal consent for interview. For the present study, we used yearly data from 1997 to 2019.

#### Analytical sample

We extracted data from available survey years between 1997 and 2020 for comparability. Our analytical sample comprised adults aged 25–64 years with complete data on weight and height and WC; pregnant or breastfeeding women were excluded. This age range was selected for comparability across studies with respect to the WHO STEPwise Approach to noncommunicable diseases (NCD) Risk Factor Surveillance (WHO-STEPS) (20).

#### Anthropometric variables

In all countries, trained personnel used standardised protocols to collect anthropometric data. Height was measured without shoes using either a fixed or portable stadiometer; and weight was measured without shoes in light clothing on a beam balance or digital weight scale. With the exception of the US, WC was measured at the mid-point between the lower edge of the rib cage and the top of the iliac crest. In the US, measurement was taken at the top of the iliac crest (See Table S1). BMI was calculated as weight in kilograms divided by height in metres squared (kg/m^2^).

We excluded participants with height <130CM or >200cm or BMI <10kg/m^2^ or >58kg/m^2^. BMI was grouped into four mutually exclusive categories as recommended by the WHO (21): underweight (<18.5kg/m^2^), normal range (18.5kg/m^2^ to 24.9kg/m^2^), overweight (25.0kg/m^2^-29.9kg/m^2^) and obesity (≥30.0kg/m^2^). Separate estimates are also provided for ≥25.0kg/m^2^, class I obesity (30.0kg/m^2^ to 34.9kg/m^2^), and class II/III obesity (≥35.0kg/m^2^). Abdominal obesity was defined as a WC ≥88cm in women or ≥102cm in men, based on the WHO and the National Heart, Lung, and Blood Institute’s/North American Association for the Study of Obesity committee recommendations (22). Participants who were outliers (<50CM or >200cm) were excluded from analyses.

### Statistical analysis

#### Descriptive analyses

Descriptive statistics for the first and last survey periods were stratified by sex and country. US data was stratified by racial-ethnic groups (collected on the basis of self-report): non-Hispanic (NH) white, NH black and Mexican-American (14). Directly age-standardised estimates were calculated using the 2000 US Census population (5-year age bands) (23).

We present the following outcomes: (i) height, weight and BMI means; (ii) BMI status; (iii) mean WC; and (iv) abdominal obesity. Secular changes in these outcomes were computed as the difference in mean/prevalence between the first and last survey periods. Sex-specific Wald tests were performed to test the null hypothesis of no change over time; the same procedure was used to test for equality of change between men and women.

#### Secular changes in BMI and WC distributions

Quantifying secular changes in the means of BMI and WC through linear regression can obscure shifts in both the location and shape of the distributions (12). Previous work (9, 12) considered BMI and WC as separate dependent variables; for comparison purposes, using all available survey years, we used quantile regression to estimate secular change (each year relative to the first) at prespecified centiles of these distributions (5^th^, 25^th^, 50^th^ (median), 75^th^, and 95^th^) (9). Survey year was treated as a categorical variable (first year as referent), and the models included continuous age and age^2^ to allow for the non-linear relation of BMI and WC to age (6). Sex-by-year interaction terms estimated differences in the magnitude of secular changes by sex.

#### Secular change in WC relative to BMI

Replicating previous analyses (9, 12), first, we used *linear regression* with BMI included as a predictor to quantify secular change in mean WC. The models included age, age^2^, BMI^2^ (to allow for the non-linear relation of WC to BMI (6, 14)) and interaction terms with survey year to estimate differences in secular changes by sex and by BMI (9). To facilitate interpretation, using the model coefficients, we quantified the sex-specific secular changes in mean WC relative to BMI at overweight and obesity cut-points (25, 30 and 35kg/m^2^). Secondly, we repeated the analysis using *quantile regression* to estimate change over time at the aforementioned WC centiles at the three BMI cut-points.

For all regression analyses, post-estimation Wald tests were used as tests of significance for men and women, and then for sex differences. All analyses accounted for each surveys’ complex design. NHANES analytical guidelines for combining consecutive 2-year cycles into 4-year periods were implemented (24). Statistical significance was set at P<0.05 for two-tailed tests, with no adjustment for multiple comparisons. Ninety-five percent confidence intervals (95% CI) are presented to convey precision. Dataset preparation and analysis were performed in SPSS V24.0 (IBM Corp., Armonk, New York) and Stata V18.0 (StataCorp, College Station, Texas). Details on the availability of survey data is provided in Table S1. All reproducible code is openly accessible via GitHub (https://github.com/shauns11/Secular-change-in-waist-circumference-relative-to-body-mass-index-).

## RESULTS

### Descriptive analyses

Tables S2A (height, weight and BMI) and S2B (WC) show the age-standardised estimates for the key anthropometric outcomes in the first and last study periods.

#### Secular change in mean BMI and WC

With the exception of Peru (both sexes), means for BMI and WC increased between the first and last survey periods in each country/US racial-ethnic group in both sexes. With regards to sex differences, mean BMI increased more in women than in men in England (1997 vs. 2019: 1.6kg/m^2^ women; 0.9kg/m^2^ men; P=0.003 for sex difference); the same pattern was observed for mean WC in Chile (2003 vs. 2017: 6.0cm women; 4.1cm men; P=0.037), England (1997 vs. 2019: 6.0cm women; 2.0cm men; P<0.001), and in US NH whites (1999-2002 vs. 2015-2018: 6.6cm women; 4.0cm men; P=0.018).

#### Secular change in overweight (including obesity), obesity and abdominal obesity

With the exceptions of Peru and US NH black women, prevalence of overweight (including obesity), obesity and abdominal obesity increased between the first and last survey periods in each country/US racial-ethnic group in both sexes.

The prevalence of overweight (including obesity) increased more in women than in men in England (1997 vs. 2019: 8.7 percentage points (pp) women; 4.4pp men; P=0.045 for sex difference) but increased more in men in US NH blacks (1999-2002 vs. 2015-2018: 11.6pp men; 3.1pp women; P=0.006). Obesity in the US increased more in men in the NH white (1999-2002 vs. 2015-2018: 14.9pp men; 8.0pp women; P=0.035) and NH black (15.9pp men; 8.4pp women; P=0.026) groups. Abdominal obesity prevalence increased more in women than in men in Chile (2003 vs. 2017: 17.6pp women; 10.4pp men; P=0.052) and in England (1997 vs. 2019: 18.4pp women; 7.7pp men; P<0.001); but increased more in men than in women in US NH blacks (1999-2002 vs. 2015-2018: 14.6pp men; 7.2pp women; P=0.027).

#### Secular change in BMI and WC distributions

Based on separate quantile regression models, the predicted values at the first and last survey periods, and the estimated secular changes (final year relative to first) at the 5^th^, 25^th^, 50^th^, 75^th^, and 95^th^ centiles, are shown in Table 1 (BMI) and Table 2 (WC). For brevity, only results for participants aged 25 years are shown (as the models contained only the main effect of age, the estimates of change are independent of age).

**Table 1.**
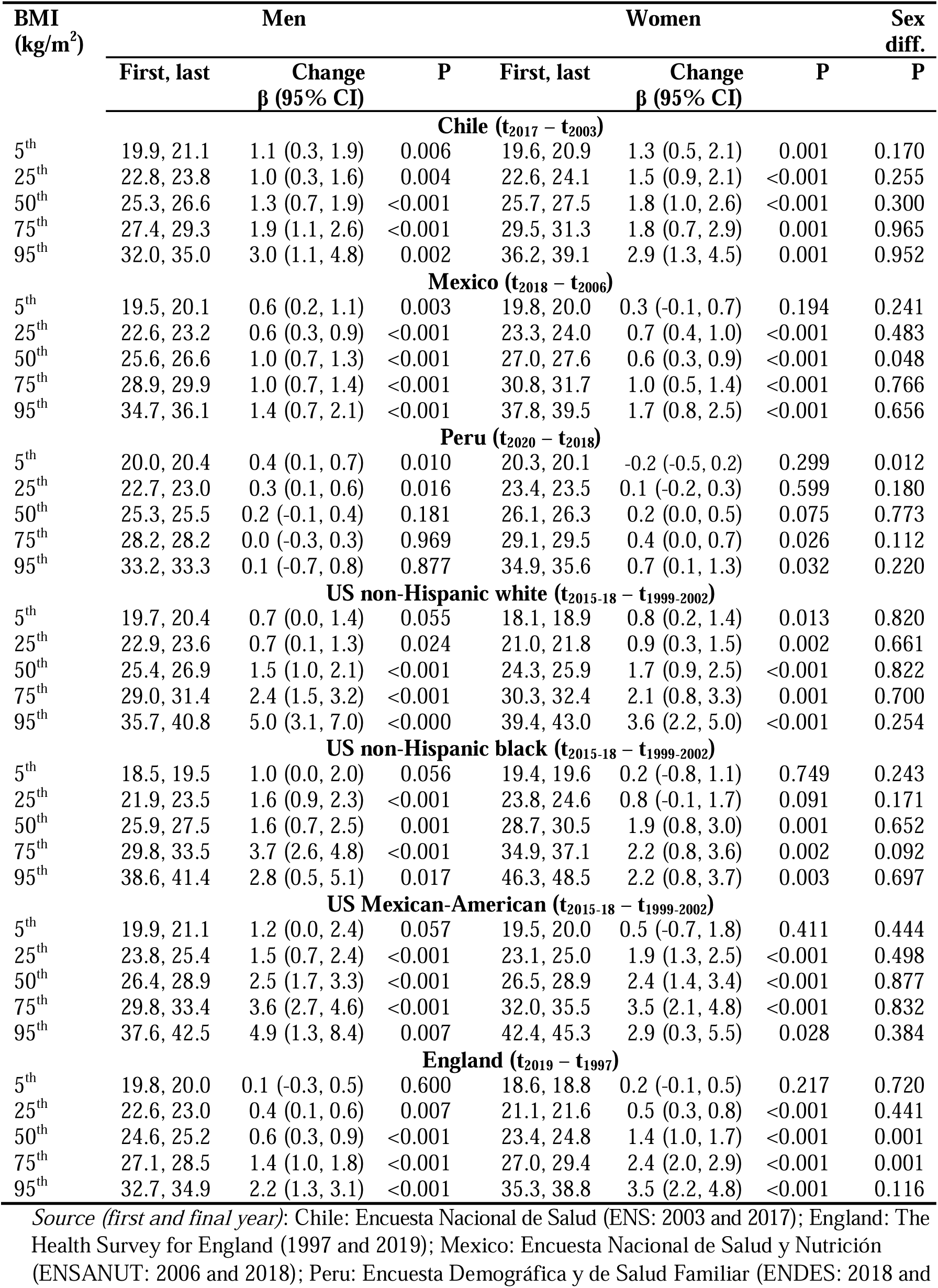
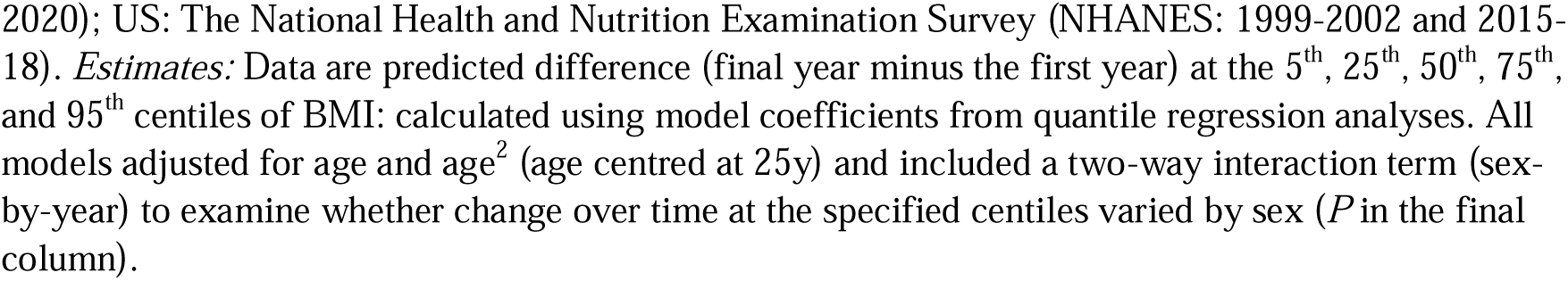
Predicted values of BMI (first and last) and difference (final minus first survey period) across centiles of BMI by sex and country.

| BMI<br>(kg/m<sup>2</sup>) | Men |  |  | Women |  |  | Sex<br>diff. |
| --- | --- | --- | --- | --- | --- | --- | --- |
| | First, last | Change<br>$\beta$ (95% CI) | P | First, last | Change<br>$\beta$ (95% CI) | P | P |
| **Chile ($t_{2017} - t_{2003}$)** |  |  |  |  |  |  |  |
| 5<sup>th</sup> | 19.9, 21.1 | 1.1 (0.3, 1.9) | 0.006 | 19.6, 20.9 | 1.3 (0.5, 2.1) | 0.001 | 0.170 |
| 25<sup>th</sup> | 22.8, 23.8 | 1.0 (0.3, 1.6) | 0.004 | 22.6, 24.1 | 1.5 (0.9, 2.1) | <0.001 | 0.255 |
| 50<sup>th</sup> | 25.3, 26.6 | 1.3 (0.7, 1.9) | <0.001 | 25.7, 27.5 | 1.8 (1.0, 2.6) | <0.001 | 0.300 |
| 75<sup>th</sup> | 27.4, 29.3 | 1.9 (1.1, 2.6) | <0.001 | 29.5, 31.3 | 1.8 (0.7, 2.9) | 0.001 | 0.965 |
| 95<sup>th</sup> | 32.0, 35.0 | 3.0 (1.1, 4.8) | 0.002 | 36.2, 39.1 | 2.9 (1.3, 4.5) | 0.001 | 0.952 |
| **Mexico ($t_{2018} - t_{2006}$)** |  |  |  |  |  |  |  |
| 5<sup>th</sup> | 19.5, 20.1 | 0.6 (0.2, 1.1) | 0.003 | 19.8, 20.0 | 0.3 (-0.1, 0.7) | 0.194 | 0.241 |
| 25<sup>th</sup> | 22.6, 23.2 | 0.6 (0.3, 0.9) | <0.001 | 23.3, 24.0 | 0.7 (0.4, 1.0) | <0.001 | 0.483 |
| 50<sup>th</sup> | 25.6, 26.6 | 1.0 (0.7, 1.3) | <0.001 | 27.0, 27.6 | 0.6 (0.3, 0.9) | <0.001 | 0.048 |
| 75<sup>th</sup> | 28.9, 29.9 | 1.0 (0.7, 1.4) | <0.001 | 30.8, 31.7 | 1.0 (0.5, 1.4) | <0.001 | 0.766 |
| 95<sup>th</sup> | 34.7, 36.1 | 1.4 (0.7, 2.1) | <0.001 | 37.8, 39.5 | 1.7 (0.8, 2.5) | <0.001 | 0.656 |
| **Peru ($t_{2020} - t_{2018}$)** |  |  |  |  |  |  |  |
| 5<sup>th</sup> | 20.0, 20.4 | 0.4 (0.1, 0.7) | 0.010 | 20.3, 20.1 | -0.2 (-0.5, 0.2) | 0.299 | 0.012 |
| 25<sup>th</sup> | 22.7, 23.0 | 0.3 (0.1, 0.6) | 0.016 | 23.4, 23.5 | 0.1 (-0.2, 0.3) | 0.599 | 0.180 |
| 50<sup>th</sup> | 25.3, 25.5 | 0.2 (-0.1, 0.4) | 0.181 | 26.1, 26.3 | 0.2 (0.0, 0.5) | 0.075 | 0.773 |
| 75<sup>th</sup> | 28.2, 28.2 | 0.0 (-0.3, 0.3) | 0.969 | 29.1, 29.5 | 0.4 (0.0, 0.7) | 0.026 | 0.112 |
| 95<sup>th</sup> | 33.2, 33.3 | 0.1 (-0.7, 0.8) | 0.877 | 34.9, 35.6 | 0.7 (0.1, 1.3) | 0.032 | 0.220 |
| **US non-Hispanic white ($t_{2015-18} - t_{1999-2002}$)** |  |  |  |  |  |  |  |
| 5<sup>th</sup> | 19.7, 20.4 | 0.7 (0.0, 1.4) | 0.055 | 18.1, 18.9 | 0.8 (0.2, 1.4) | 0.013 | 0.820 |
| 25<sup>th</sup> | 22.9, 23.6 | 0.7 (0.1, 1.3) | 0.024 | 21.0, 21.8 | 0.9 (0.3, 1.5) | 0.002 | 0.661 |
| 50<sup>th</sup> | 25.4, 26.9 | 1.5 (1.0, 2.1) | <0.001 | 24.3, 25.9 | 1.7 (0.9, 2.5) | <0.001 | 0.822 |
| 75<sup>th</sup> | 29.0, 31.4 | 2.4 (1.5, 3.2) | <0.001 | 30.3, 32.4 | 2.1 (0.8, 3.3) | 0.001 | 0.700 |
| 95<sup>th</sup> | 35.7, 40.8 | 5.0 (3.1, 7.0) | <0.000 | 39.4, 43.0 | 3.6 (2.2, 5.0) | <0.001 | 0.254 |
| **US non-Hispanic black ($t_{2015-18} - t_{1999-2002}$)** |  |  |  |  |  |  |  |
| 5<sup>th</sup> | 18.5, 19.5 | 1.0 (0.0, 2.0) | 0.056 | 19.4, 19.6 | 0.2 (-0.8, 1.1) | 0.749 | 0.243 |
| 25<sup>th</sup> | 21.9, 23.5 | 1.6 (0.9, 2.3) | <0.001 | 23.8, 24.6 | 0.8 (-0.1, 1.7) | 0.091 | 0.171 |
| 50<sup>th</sup> | 25.9, 27.5 | 1.6 (0.7, 2.5) | 0.001 | 28.7, 30.5 | 1.9 (0.8, 3.0) | 0.001 | 0.652 |
| 75<sup>th</sup> | 29.8, 33.5 | 3.7 (2.6, 4.8) | <0.001 | 34.9, 37.1 | 2.2 (0.8, 3.6) | 0.002 | 0.092 |
| 95<sup>th</sup> | 38.6, 41.4 | 2.8 (0.5, 5.1) | 0.017 | 46.3, 48.5 | 2.2 (0.8, 3.7) | 0.003 | 0.697 |
| **US Mexican-American ($t_{2015-18} - t_{1999-2002}$)** |  |  |  |  |  |  |  |
| 5<sup>th</sup> | 19.9, 21.1 | 1.2 (0.0, 2.4) | 0.057 | 19.5, 20.0 | 0.5 (-0.7, 1.8) | 0.411 | 0.444 |
| 25<sup>th</sup> | 23.8, 25.4 | 1.5 (0.7, 2.4) | <0.001 | 23.1, 25.0 | 1.9 (1.3, 2.5) | <0.001 | 0.498 |
| 50<sup>th</sup> | 26.4, 28.9 | 2.5 (1.7, 3.3) | <0.001 | 26.5, 28.9 | 2.4 (1.4, 3.4) | <0.001 | 0.877 |
| 75<sup>th</sup> | 29.8, 33.4 | 3.6 (2.7, 4.6) | <0.001 | 32.0, 35.5 | 3.5 (2.1, 4.8) | <0.001 | 0.832 |
| 95<sup>th</sup> | 37.6, 42.5 | 4.9 (1.3, 8.4) | 0.007 | 42.4, 45.3 | 2.9 (0.3, 5.5) | 0.028 | 0.384 |
| **England ($t_{2019} - t_{1997}$)** |  |  |  |  |  |  |  |
| 5<sup>th</sup> | 19.8, 20.0 | 0.1 (-0.3, 0.5) | 0.600 | 18.6, 18.8 | 0.2 (-0.1, 0.5) | 0.217 | 0.720 |
| 25<sup>th</sup> | 22.6, 23.0 | 0.4 (0.1, 0.6) | 0.007 | 21.1, 21.6 | 0.5 (0.3, 0.8) | <0.001 | 0.441 |
| 50<sup>th</sup> | 24.6, 25.2 | 0.6 (0.3, 0.9) | <0.001 | 23.4, 24.8 | 1.4 (1.0, 1.7) | <0.001 | 0.001 |
| 75<sup>th</sup> | 27.1, 28.5 | 1.4 (1.0, 1.8) | <0.001 | 27.0, 29.4 | 2.4 (2.0, 2.9) | <0.001 | 0.001 |
| 95<sup>th</sup> | 32.7, 34.9 | 2.2 (1.3, 3.1) | <0.001 | 35.3, 38.8 | 3.5 (2.2, 4.8) | <0.001 | 0.116 |

Source (first and final year): Chile: Encuesta Nacional de Salud (ENS: 2003 and 2017); England: The Health Survey for England (1997 and 2019); Mexico: Encuesta Nacional de Salud y Nutrición (ENSANUT: 2006 and 2018); Peru: Encuesta Demográfica y de Salud Familiar (ENDES: 2018 and
2020); US: The National Health and Nutrition Examination Survey (NHANES: 1999-2002 and 2015-18). *Estimates*: Data are predicted difference (final year minus the first year) at the 5<sup>th</sup>, 25<sup>th</sup>, 50<sup>th</sup>, 75<sup>th</sup>, and 95<sup>th</sup> centiles of BMI: calculated using model coefficients from quantile regression analyses. All models adjusted for age and age<sup>2</sup> (age centred at 25y) and included a two-way interaction term (sex-by-year) to examine whether change over time at the specified centiles varied by sex (*P* in the final column).

**Table 2.**
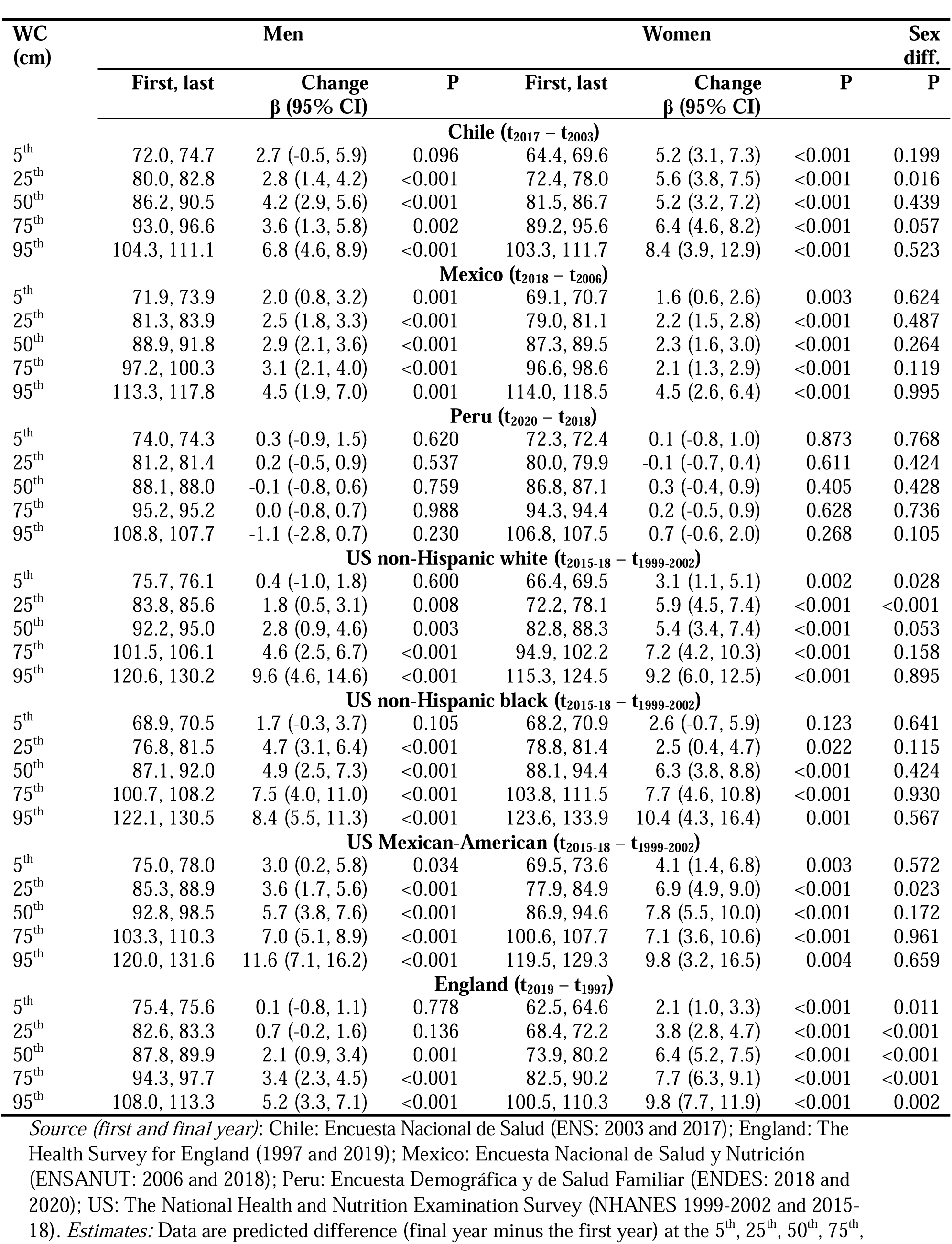
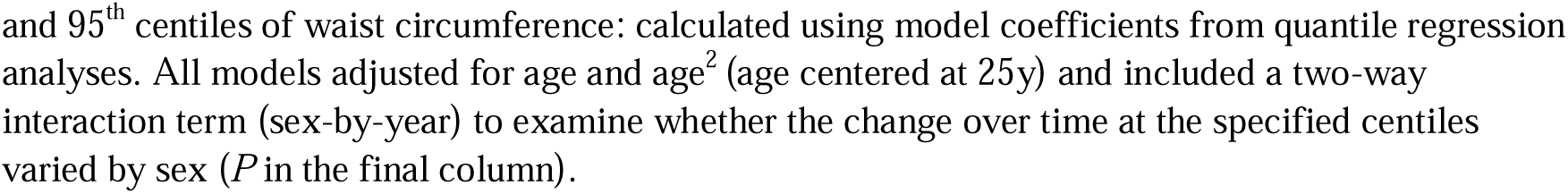
Predicted values of waist circumference (first and last) and difference (final minus first survey period) across centiles of waist circumference by sex and country.

| WC<br>(cm) | Men |  |  | Women |  |  | Sex<br>diff. |
| --- | --- | --- | --- | --- | --- | --- | --- |
| | First, last | Change<br>$\beta$ (95% CI) | P | First, last | Change<br>$\beta$ (95% CI) | P | P |
| **Chile ($t_{2017} - t_{2003}$)** |  |  |  |  |  |  |  |
| 5<sup>th</sup> | 72.0, 74.7 | 2.7 (-0.5, 5.9) | 0.096 | 64.4, 69.6 | 5.2 (3.1, 7.3) | <0.001 | 0.199 |
| 25<sup>th</sup> | 80.0, 82.8 | 2.8 (1.4, 4.2) | <0.001 | 72.4, 78.0 | 5.6 (3.8, 7.5) | <0.001 | 0.016 |
| 50<sup>th</sup> | 86.2, 90.5 | 4.2 (2.9, 5.6) | <0.001 | 81.5, 86.7 | 5.2 (3.2, 7.2) | <0.001 | 0.439 |
| 75<sup>th</sup> | 93.0, 96.6 | 3.6 (1.3, 5.8) | 0.002 | 89.2, 95.6 | 6.4 (4.6, 8.2) | <0.001 | 0.057 |
| 95<sup>th</sup> | 104.3, 111.1 | 6.8 (4.6, 8.9) | <0.001 | 103.3, 111.7 | 8.4 (3.9, 12.9) | <0.001 | 0.523 |
| **Mexico ($t_{2018} - t_{2006}$)** |  |  |  |  |  |  |  |
| 5<sup>th</sup> | 71.9, 73.9 | 2.0 (0.8, 3.2) | 0.001 | 69.1, 70.7 | 1.6 (0.6, 2.6) | 0.003 | 0.624 |
| 25<sup>th</sup> | 81.3, 83.9 | 2.5 (1.8, 3.3) | <0.001 | 79.0, 81.1 | 2.2 (1.5, 2.8) | <0.001 | 0.487 |
| 50<sup>th</sup> | 88.9, 91.8 | 2.9 (2.1, 3.6) | <0.001 | 87.3, 89.5 | 2.3 (1.6, 3.0) | <0.001 | 0.264 |
| 75<sup>th</sup> | 97.2, 100.3 | 3.1 (2.1, 4.0) | <0.001 | 96.6, 98.6 | 2.1 (1.3, 2.9) | <0.001 | 0.119 |
| 95<sup>th</sup> | 113.3, 117.8 | 4.5 (1.9, 7.0) | 0.001 | 114.0, 118.5 | 4.5 (2.6, 6.4) | <0.001 | 0.995 |
| **Peru ($t_{2020} - t_{2018}$)** |  |  |  |  |  |  |  |
| 5<sup>th</sup> | 74.0, 74.3 | 0.3 (-0.9, 1.5) | 0.620 | 72.3, 72.4 | 0.1 (-0.8, 1.0) | 0.873 | 0.768 |
| 25<sup>th</sup> | 81.2, 81.4 | 0.2 (-0.5, 0.9) | 0.537 | 80.0, 79.9 | -0.1 (-0.7, 0.4) | 0.611 | 0.424 |
| 50<sup>th</sup> | 88.1, 88.0 | -0.1 (-0.8, 0.6) | 0.759 | 86.8, 87.1 | 0.3 (-0.4, 0.9) | 0.405 | 0.428 |
| 75<sup>th</sup> | 95.2, 95.2 | 0.0 (-0.8, 0.7) | 0.988 | 94.3, 94.4 | 0.2 (-0.5, 0.9) | 0.628 | 0.736 |
| 95<sup>th</sup> | 108.8, 107.7 | -1.1 (-2.8, 0.7) | 0.230 | 106.8, 107.5 | 0.7 (-0.6, 2.0) | 0.268 | 0.105 |
| **US non-Hispanic white ($t_{2015-18} - t_{1999-2002}$)** |  |  |  |  |  |  |  |
| 5<sup>th</sup> | 75.7, 76.1 | 0.4 (-1.0, 1.8) | 0.600 | 66.4, 69.5 | 3.1 (1.1, 5.1) | 0.002 | 0.028 |
| 25<sup>th</sup> | 83.8, 85.6 | 1.8 (0.5, 3.1) | 0.008 | 72.2, 78.1 | 5.9 (4.5, 7.4) | <0.001 | <0.001 |
| 50<sup>th</sup> | 92.2, 95.0 | 2.8 (0.9, 4.6) | 0.003 | 82.8, 88.3 | 5.4 (3.4, 7.4) | <0.001 | 0.053 |
| 75<sup>th</sup> | 101.5, 106.1 | 4.6 (2.5, 6.7) | <0.001 | 94.9, 102.2 | 7.2 (4.2, 10.3) | <0.001 | 0.158 |
| 95<sup>th</sup> | 120.6, 130.2 | 9.6 (4.6, 14.6) | <0.001 | 115.3, 124.5 | 9.2 (6.0, 12.5) | <0.001 | 0.895 |
| **US non-Hispanic black ($t_{2015-18} - t_{1999-2002}$)** |  |  |  |  |  |  |  |
| 5<sup>th</sup> | 68.9, 70.5 | 1.7 (-0.3, 3.7) | 0.105 | 68.2, 70.9 | 2.6 (-0.7, 5.9) | 0.123 | 0.641 |
| 25<sup>th</sup> | 76.8, 81.5 | 4.7 (3.1, 6.4) | <0.001 | 78.8, 81.4 | 2.5 (0.4, 4.7) | 0.022 | 0.115 |
| 50<sup>th</sup> | 87.1, 92.0 | 4.9 (2.5, 7.3) | <0.001 | 88.1, 94.4 | 6.3 (3.8, 8.8) | <0.001 | 0.424 |
| 75<sup>th</sup> | 100.7, 108.2 | 7.5 (4.0, 11.0) | <0.001 | 103.8, 111.5 | 7.7 (4.6, 10.8) | <0.001 | 0.930 |
| 95<sup>th</sup> | 122.1, 130.5 | 8.4 (5.5, 11.3) | <0.001 | 123.6, 133.9 | 10.4 (4.3, 16.4) | 0.001 | 0.567 |
| **US Mexican-American ($t_{2015-18} - t_{1999-2002}$)** |  |  |  |  |  |  |  |
| 5<sup>th</sup> | 75.0, 78.0 | 3.0 (0.2, 5.8) | 0.034 | 69.5, 73.6 | 4.1 (1.4, 6.8) | 0.003 | 0.572 |
| 25<sup>th</sup> | 85.3, 88.9 | 3.6 (1.7, 5.6) | <0.001 | 77.9, 84.9 | 6.9 (4.9, 9.0) | <0.001 | 0.023 |
| 50<sup>th</sup> | 92.8, 98.5 | 5.7 (3.8, 7.6) | <0.001 | 86.9, 94.6 | 7.8 (5.5, 10.0) | <0.001 | 0.172 |
| 75<sup>th</sup> | 103.3, 110.3 | 7.0 (5.1, 8.9) | <0.001 | 100.6, 107.7 | 7.1 (3.6, 10.6) | <0.001 | 0.961 |
| 95<sup>th</sup> | 120.0, 131.6 | 11.6 (7.1, 16.2) | <0.001 | 119.5, 129.3 | 9.8 (3.2, 16.5) | 0.004 | 0.659 |
| **England ($t_{2019} - t_{1997}$)** |  |  |  |  |  |  |  |
| 5<sup>th</sup> | 75.4, 75.6 | 0.1 (-0.8, 1.1) | 0.778 | 62.5, 64.6 | 2.1 (1.0, 3.3) | <0.001 | 0.011 |
| 25<sup>th</sup> | 82.6, 83.3 | 0.7 (-0.2, 1.6) | 0.136 | 68.4, 72.2 | 3.8 (2.8, 4.7) | <0.001 | <0.001 |
| 50<sup>th</sup> | 87.8, 89.9 | 2.1 (0.9, 3.4) | 0.001 | 73.9, 80.2 | 6.4 (5.2, 7.5) | <0.001 | <0.001 |
| 75<sup>th</sup> | 94.3, 97.7 | 3.4 (2.3, 4.5) | <0.001 | 82.5, 90.2 | 7.7 (6.3, 9.1) | <0.001 | <0.001 |
| 95<sup>th</sup> | 108.0, 113.3 | 5.2 (3.3, 7.1) | <0.001 | 100.5, 110.3 | 9.8 (7.7, 11.9) | <0.001 | 0.002 |

*Source (first and final year):* Chile: Encuesta Nacional de Salud (ENS: 2003 and 2017); England: The Health Survey for England (1997 and 2019); Mexico: Encuesta Nacional de Salud y Nutrición (ENSANUT: 2006 and 2018); Peru: Encuesta Demográfica y de Salud Familiar (ENDES: 2018 and 2020); US: The National Health and Nutrition Examination Survey (NHANES 1999-2002 and 2015-18). *Estimates:* Data are predicted difference (final year minus the first year) at the 5<sup>th</sup>, 25<sup>th</sup>, 50<sup>th</sup>, 75<sup>th</sup>,
and 95<sup>th</sup> centiles of waist circumference: calculated using model coefficients from quantile regression analyses. All models adjusted for age and age<sup>2</sup> (age centered at 25y) and included a two-way interaction term (sex-by-year) to examine whether the change over time at the specified centiles varied by sex (*P* in the final column).

BMI (except men in Peru) and WC (except both sexes in Peru) showed larger increases at the upper-tails of the distribution in both sexes. Significant differences by sex at the upper-tail were evident only in England. Increases between 1997 and 2019 at the 50^th^ and 75^th^ BMI centiles were larger in women than in men (e.g., 50^th^ centile: 1.4kg/m^2^ women; 0.6kg/m^2^ men: P=0.001 for sex difference); over the same time period, WC at the 50^th^ centile increased by 6.4cm and 2.1cm in women and men, respectively (P<0.001 for sex difference).

#### Secular change in WC relative to BMI

Based on the model coefficients with BMI included as an additional predictor of WC, the sex-specific secular changes (final year relative to the first) in mean WC (linear regressions) and at the 5^th^, 25^th^, 50^th^, 75^th^, and 95^th^ centiles (quantile regressions) at BMI = 25kg/m^2^, 30kg/m^2^ and 35kg/m^2^ are shown in Figures 1-5. Full estimates are provided in Tables S3A-S3G.

**Figure 1.**
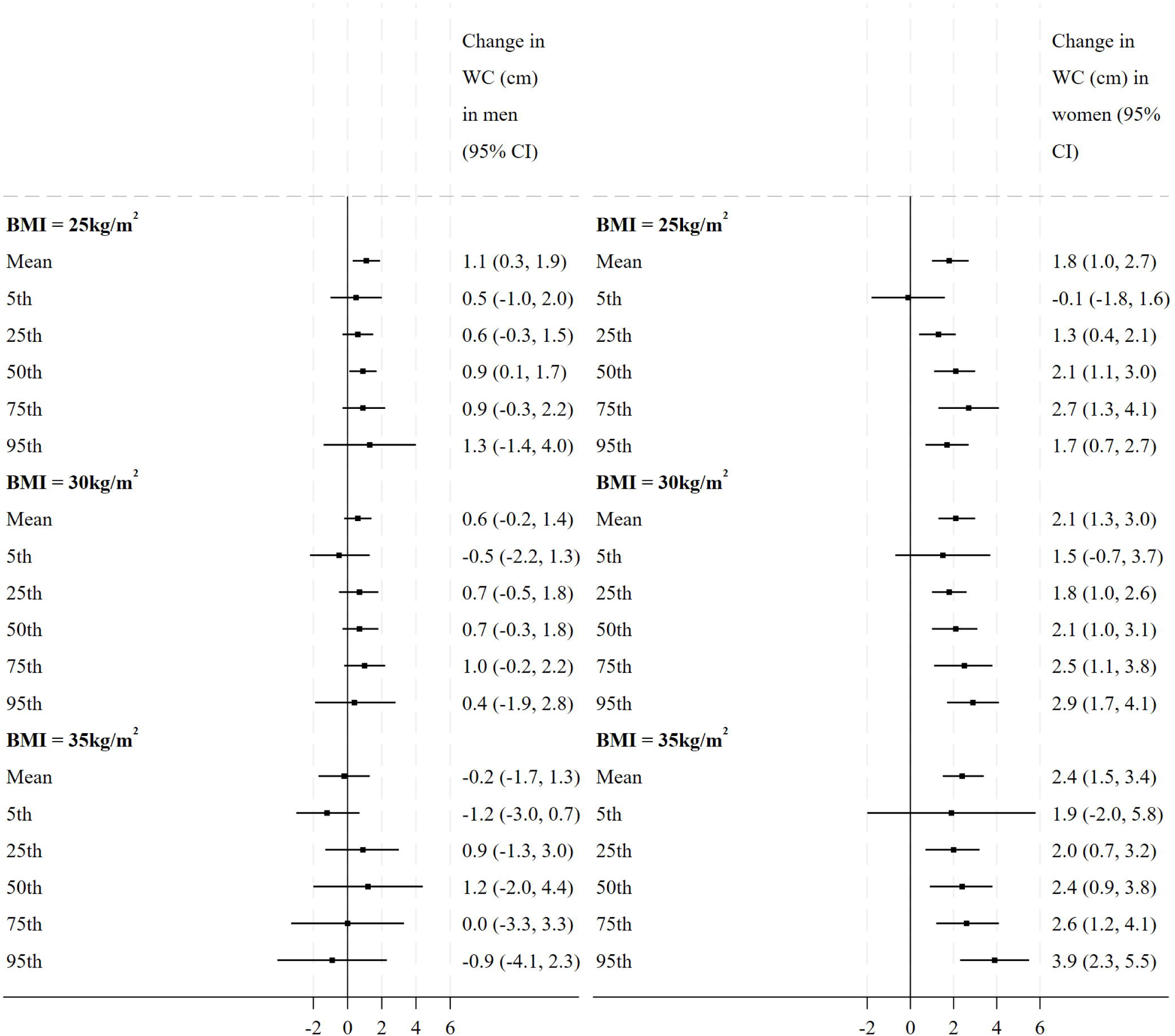
Estimated secular change (2003 to 2017) in mean WC and at various centiles of the WC distribution relative to BMI at 25, 30 and 35kg/m^2^ by sex in Chile

**Figure 2.**
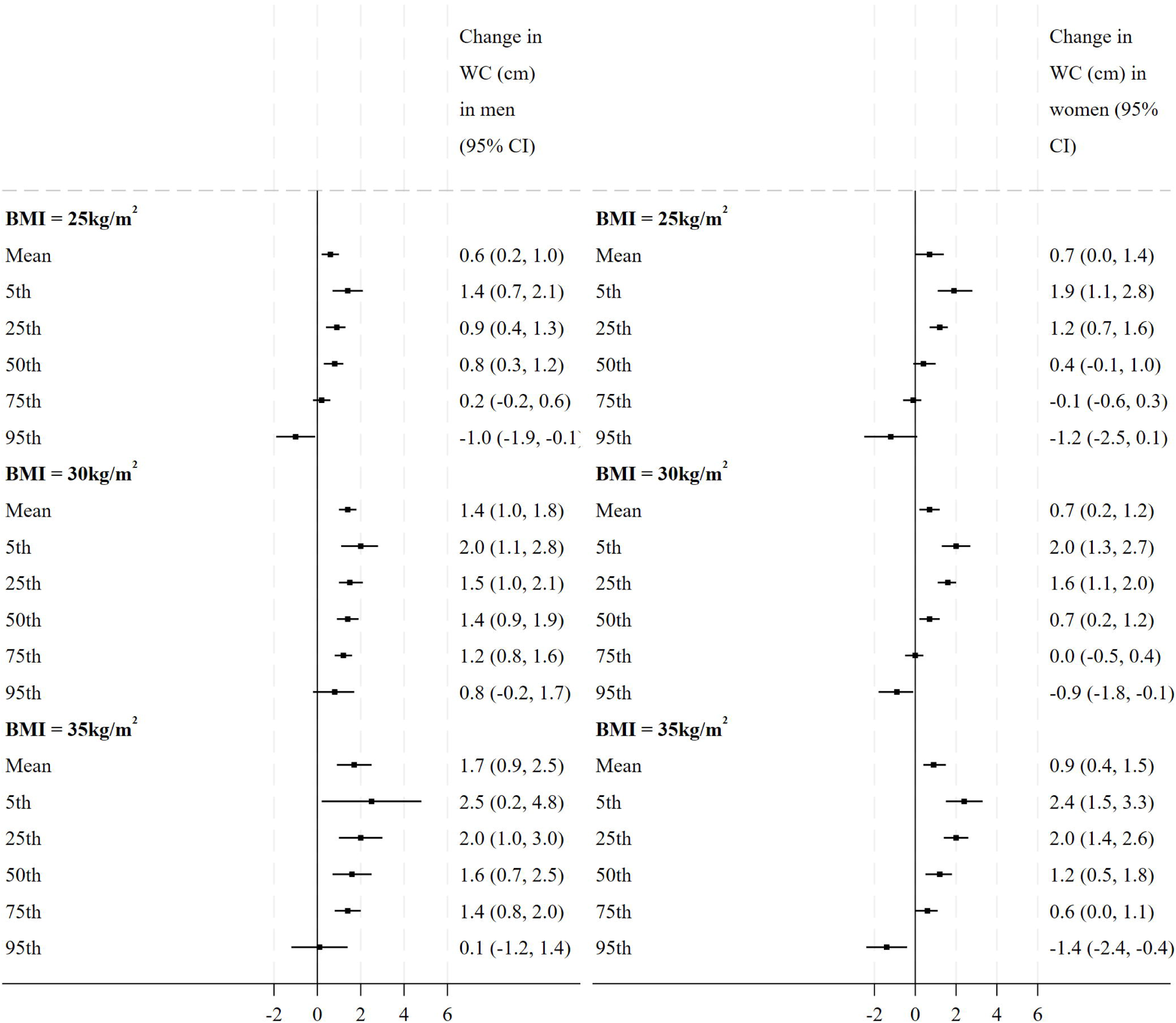
Estimated secular change (2006 to 2018) in mean WC and at various centiles of the WC distribution relative to BMI at 25, 30 and 35kg/m^2^ by sex in Mexico

**Figure 3.**
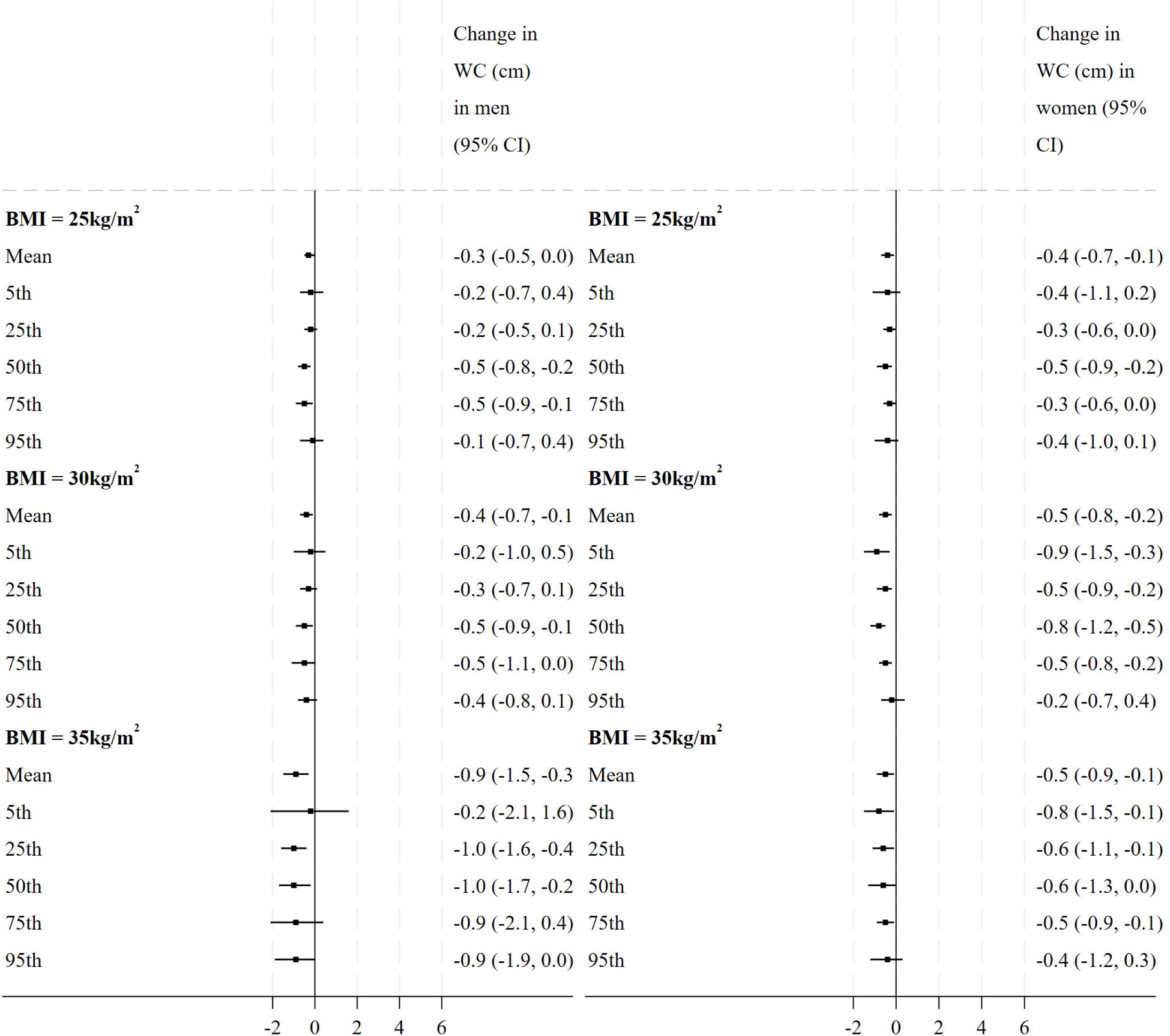
Estimated secular change (2018 to 2020) in mean WC and at various centiles of the WC distribution relative to BMI at 25, 30 and 35kg/m^2^ by sex in Peru

**Figure 4.**
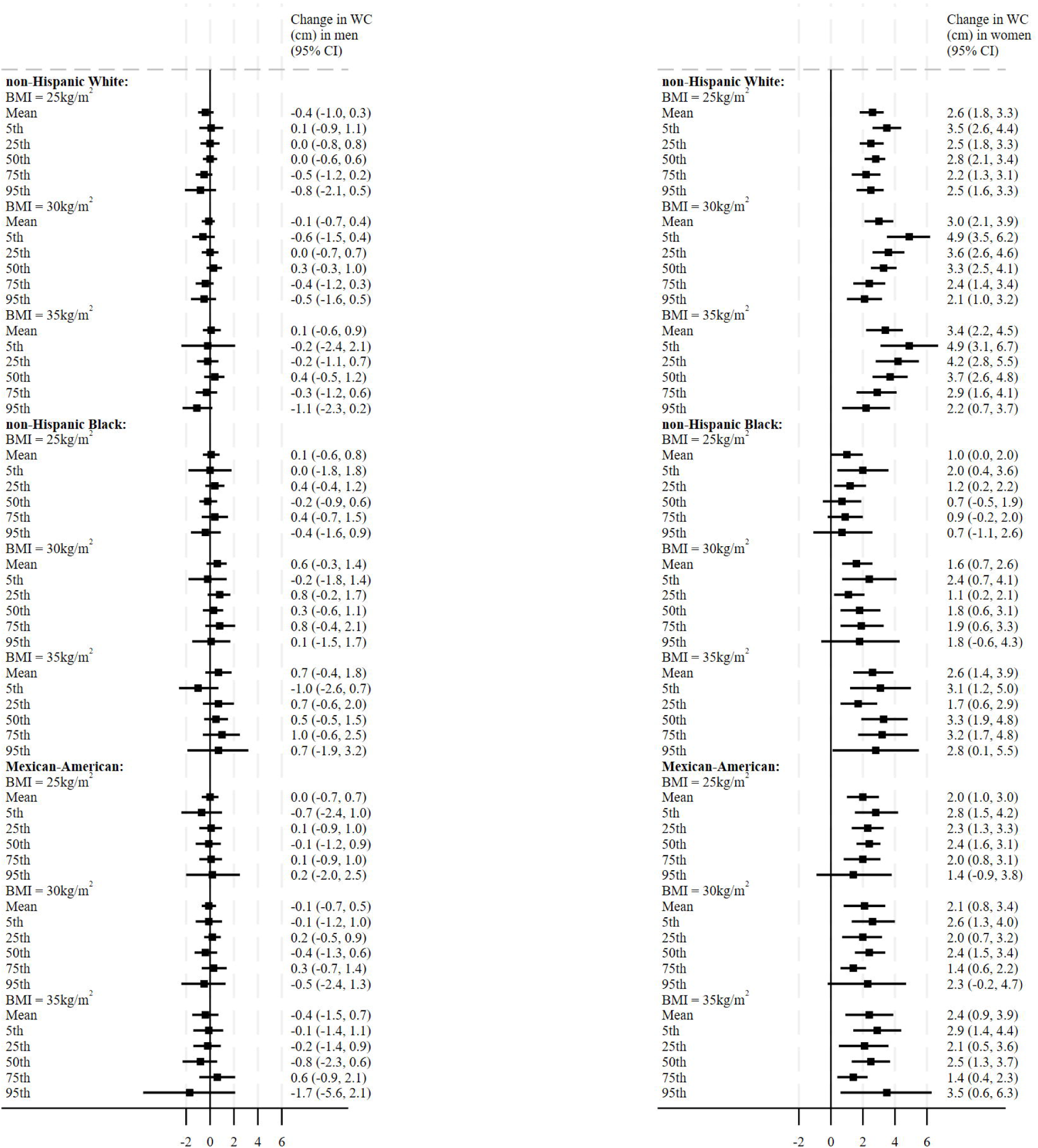
Estimated secular change (1999-2002 to 2015-18) in mean WC and at various centiles of the WC distribution relative to BMI at 25, 30 and 35kg/m^2^ by sex and racial-ethnic group in the US

**Figure 5.**
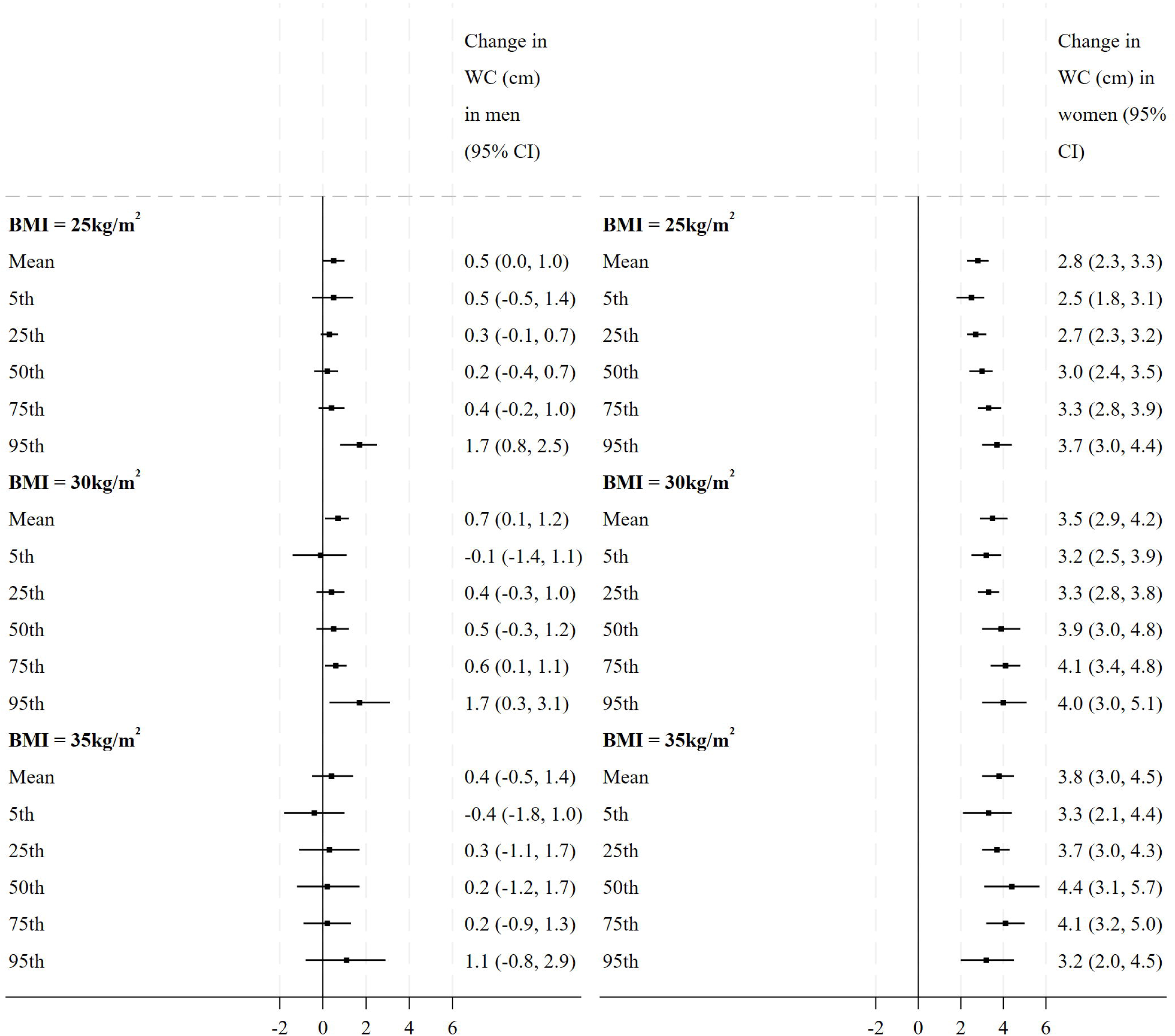
Estimated secular change (1999 to 2017) in mean WC and at various centiles of the WC distribution relative to BMI at 25, 30 and 35kg/m^2^ by sex in England

#### Linear regression: change in mean WC relative to BMI

Mixed patterns were observed for secular changes in mean WC relative to BMI. Mean WC decreased between 2018 and 2020 at each BMI level in both sexes in Peru.

In men, mean WC increased at each BMI level in Mexico, at BMI = 25kg/m^2^ in Chile, and at BMI = 30kg/m^2^ in England. In women, mean WC increased at each BMI level in Chile, Mexico, England and in each US racial-ethnic group. Increases in mean WC were higher at higher BMI levels. For example, at BMI = 25kg/m^2^, increases in mean WC ranged from 0.7cm in Mexico (2006 vs. 2018; 95% CI: 0.0-1.4) to 2.8cm in England (1997 vs. 2019; 95% CI: 2.3-3.3). At BMI = 35kg/m^2^, increases in mean WC ranged from 0.9cm in Mexico (2006 vs. 2018; 95% CI: 0.4-1.5) to 3.8cm in England (1997 vs. 2019; 95% CI: 3.0-4.5).

Sex differences in secular changes in mean WC relative to BMI were observed in US NH whites, US Mexican-Americans, and England, with larger increases in women than in men at each BMI cut-point.

#### Quantile regressions: change in WC distributions relative to BMI

Mixed patterns were observed for secular changes across the distributions of WC relative to BMI. Here we discuss only the results at the 50^th^ and 75^th^ WC centiles at BMI = 25kg/m^2^ and 30kg/m^2^, and present P-values where change over time varied by sex. Complete results are shown in Tables S3A-S3G.

In both sexes in Peru, WC at the 50^th^ and 75^th^ centiles decreased between 2018 and 2020 at both BMI cut-points. In men, at BMI = 25kg/m^2^, increases at the 50^th^ WC centile were observed in Chile and in Mexico. At BMI = 30kg/m^2^, increases at the 50^th^ and 75^th^ WC centiles were observed in Mexico and in England.

Increases in WC relative to BMI were more evident in women, with larger increases at higher levels of BMI, especially in US NH whites and in England (Figures 4 and 5, respectively). At BMI = 25kg/m^2^, increases at the 50^th^ WC centile ranged from 2.3cm in US Mexican-Americans (1999-2002 vs. 2015-2018; 95% CI: 1.6-3.1; P<0.001 for sex difference) to 3.0cm in England (1997 vs. 2019; 95% CI: 2.4-3.5; P<0.001). Increases at the 75^th^ WC centile ranged from 1.9cm in US Mexican-Americans (95% CI: 0.8-3.1; P=0.013) to 3.3cm in England (95% CI: 2.8-3.9; P<0.001). At BMI = 30kg/m^2^, increases at the 50^th^ WC centile ranged from 0.7cm in Mexico (2006 vs. 2018; 95% CI: 0.2-1.2; P=0.045 for sex difference, but with a larger increase in men) to 3.9cm in England (1997 vs. 2019; 95% CI: 3.0-4.8; P<0.001). Increases at the 75^th^ WC centile ranged from 1.4cm in US Mexican-Americans (1999-2002 vs. 2015-2018; 95% CI: 0.6-2.2) to 4.1cm in England (1997 vs. 2019; 95% CI: 3.4-4.8; P<0.001).

## DISCUSSION

Using nationally-representative HES data from US, Mexico, Chile, Peru and England, we quantified secular changes between 1997 and 2020 in BMI, WC, and WC relative to BMI, and explored differences by sex.

We observed that, except for Peru, average levels of BMI and WC increased between the first and last survey periods in each country/US racial-ethnic group in both sexes, with larger increases in women than in men in England (BMI and WC), Chile (WC) and US NH whites (WC). Except for Peru, BMI and WC each showed an upward shift in each country/US racial-ethnic group in both sexes, with larger increases at the upper-tails. Increases in BMI and WC at the 50^th^ and 75^th^ centiles were larger in women than in men in England.

Secular changes in WC relative to BMI showed mixed patterns. Decreases in mean and median WC relative to BMI between 2018 and 2020 were observed in both sexes in Peru. Increases over time at the 50^th^ and 75^th^ WC centiles relative to BMI were more pronounced in women than in men, with larger increases at higher levels of BMI especially in US NH whites and in England. Our analyses showed that Mexico was a noteworthy exception: secular change in WC relative to BMI was larger in men than in women.

### Comparisons with other studies

Precise comparisons with the few global studies that have examined secular changes in mean WC relative to BMI (9, 14) or to body weight (13) are difficult due to differences in study populations, time period, analytical sample (including age range and sex), and statistical techniques. Nevertheless, our findings agree with previous studies which have shown increases in WC in both sexes and the shift in recent years to higher WC independently of BMI, especially, but not exclusively, in women. Most of the secular increase in mean WC between 1999-2000 and 2011-2012 in the US was independent of increases in BMI in women but not in men, with no reported differences between racial-ethnic groups (6). Albrecht et al. reported disproportionate increases in mean WC relative to BMI in US, Mexico, China, and England populations, particularly in women aged 20-29 years (14).

Our study updates and expands on these findings. For example, our model-based predictions for the final survey period suggest that the median WC in women aged 25 years in Mexico and in each US racial-ethnic group exceeds the 88cm threshold used to define abdominal obesity (Table 2).

### Public health implications

These recent secular increases in WC relative to BMI in Chile, Mexico, US and England highlight that future disease risk, such as those associated with the amount of visceral fat, is potentially underestimated by relying on BMI alone (6). Such increases in WC may partly explain concomitant increases in diabetes and prediabetes among individuals with class II/III obesity in the US (25) and metabolic syndrome in Asians (26). Factors such as higher sedentary activity, sleep deprivation, diets high in sugar and refined carbohydrates/energy dense foods, endocrine disruptors, and certain medications have been proposed in the literature as potential explanations for the recent increases in WC relative to BMI (27, 28). Speculative reasons for the greater increase in WC relative to BMI in women than in men include higher levels of percent body fat (29) and physiological differences pronounced at older ages (mainly after menopause), as the deposits of subcutaneous adipose tissue surrounding hips and thighs decrease with increasing age, and visceral adipose tissue increases (30).

Sex differences in the secular changes in WC relative to BMI are important to consider as WC is a major indicator of cardiovascular and metabolic risk, and has been shown to be more strongly associated with the risk of adverse health outcomes than BMI, especially in women (31). In a Consensus Statement, the International Atherosclerosis Society and International Chair on Cardiometabolic Risk Working Group on Visceral Obesity recommended that WC be included routinely in clinical practice (8).

### Strengths and limitations

The five countries analysed herein allowed examination of secular changes in WC relative to BMI across countries with different levels of income (*High-income*: Chile, England, US; *Upper-Middle income*: Mexico; *Middle-income*: Peru), but with a focus on the Americas region. Our study adds to the global evidence base by adding the most recently available HES data from England and the US, and including data, for the first time, from Chile and Peru, and from men in Mexico. Data was extracted from each country’s national HES, enabling nationally-representative inference with high-quality data: height, weight and WC were directly measured by trained study staff using standardised protocols, and so our findings avoid the known biases associated with self-reported measures (32). The time period covered spanned more than a decade in four of the five countries; this allowed us to account for the potential effects of any obesity prevention-related policies implemented during this time. Further we used all available data to produce model-based estimates of secular change, and employed linear and quantile regression to examine changes over time in means and across the WC distributions independently of BMI.

Several limitations should be considered in the interpretation of our results. Our study made no attempt to directly compare the magnitude of secular changes across countries due to differences in survey periods. Instead, we highlight similarities and differences in overall patterns, including differences between men and women. Although WC measurement was consistent within each HES, comparisons between the US and the other countries should be additionally treated with caution as the former measured WC at the top of the iliac crest rather than at the mid-point between the iliac crest and the costal wall. There is some evidence to suggest WC measurements at the top of the iliac crest versus the mid-point are larger, especially in women (33–34). Peruvian data could only be presented from 2018 to 2020, hence these findings on secular changes should be treated with caution. Survey non-response is an increasing problem. Participants in health examination surveys tend to be healthier than non-participants. Furthermore, conditional on survey participation (e.g. data on socio-demographics), less healthy participants, including groups with higher BMI, may be less likely to agree to direct measurements of height, weight and WC (6, 32). Although non-response weights are available (and were used herein), such weights do not necessarily adjust for potential bias in anthropometric data collection. Our findings may therefore have underestimated the secular increases in WC relative to BMI to some extent. Combining adjacent 2-year cycles in NHANES to produce estimates based on four years of data ensured greater precision and smaller sampling error; however, doing so makes the inherent assumption of no trend in the estimate over the time period being combined (24). Finally, apart from NHANES, the other HES do not routinely collect body composition data, and so anthropometric indicators potentially more sensitive to adiposity (e.g., total or percentage body fat, or skinfold thickness) were not available.

## CONCLUSION

Our analyses of twenty years of anthropometric data support emerging evidence of increasing WC relative to BMI, especially, but not exclusively, in women. Routinely collecting WC data in health surveys strengthens national efforts of disease risk surveillance and potentially contributes to reduced disparities across BMI groups for detection and disease treatment. More investigation is needed to identify the key modifiable factors underlying recent increases in WC relative to BMI.

## Abbreviations

BMI: body mass index
cm: centimetres
ENDES: Encuesta Demográfica y de Salud Familiar (Peruvian Health Survey)
ENS: Encuesta Nacional de Salud (Chilean Health Survey)
ENSANUT: Encuesta Nacional de Salud y Nutrición (Mexican Health Survey)
HES: health examination surveys
HSE: Health Survey for England
NH: non-Hispanic
NHANES: National Health and Nutrition Examination Survey
PP: percentage points
WC: waist circumference
WHO: World Health Organization

## Availability of data and materials

Source data were openly available before the initiation of the study. *Chile:* National health survey databases are freely available on the Ministry of Health website (http://epi.minsal.cl/bases-de-datos). *Mexico*: The data underlying this study were generated by the National Institute of Public Health, and are freely available (https://ensanut.insp.mx/). *Peru*: The data underlying this study can be found in the repository National Open Data Plataform (https://www.datosabiertos.gob.pe/) and are freely accessible. *US*: NHANES datasets are publicly available at the CDC website (https://www.cdc.gov/nchs/nhanes/index.htm) *England:* The Health Survey for England datasets generated and analysed during the current study are available via the UK Data Service (UKDS: https://ukdataservice.ac.uk/), subject to their End-user Licence.

## Data Availability

Source data were openly available before the initiation of the study. Chile: National health survey databases are freely available on the Ministry of Health website (http://epi.minsal.cl/bases-de-datos). Mexico: The data underlying this study were generated by the National Institute of Public Health, and are freely available (https://ensanut.insp.mx/). Peru: The data underlying this study can be found in the repository National Open Data Plataform (https://www.datosabiertos.gob.pe/) and are freely accessible. US: NHANES datasets are publicly available at the CDC website (https://www.cdc.gov/nchs/nhanes/index.htm) England: The Health Survey for England datasets generated and analysed during the current study are available via the UK Data Service (UKDS: https://ukdataservice.ac.uk/), subject to their End-user Licence.

## Acknowledgements

The authors thank the interviewers and nurses, and participants in the five national health surveys. The authors also thank the leading agencies (Chile: the Pontificia Universidad Católica de Chile (PUC) and the Ministry of Health); Mexico: Instituto Nacional de Nutrición (INSP); Peru: the National Institute of Statistics and Informatics; US: National Center for Health Statistics at the Centers for Disease Control and Prevention; England: NatCen Social Research, and NHS England) responsible for designing, collecting and administering the survey data and making the data available for public use. JM and SS also thank colleagues at NatCen Social Research, and NHS England.

## Authors Contributions

LMSR and SS conceptualised the study. SS, JSM and AP were responsible for collecting information, generating the data sets, and conducting the analyses. LMSR and SS interpreted the results and wrote the manuscript. JM, ABO, LTM and AM critically revised the manuscript. All authors have read and approved the manuscript.

