## Supplementary material for "Sex differences in the secular change in waist circumference relative to body mass index in the Americas and England from 1997 to 2020"

**Table S1** Description of the design and methods of the national health surveys for each of the five countries.

| **Country** | **Chile[1]** | **Mexico[2]** | **Peru[3]** | **United States[4]** | **England[5]** |
|---|---|---|---|---|---|
| **Survey** | **ENS** | **ENSANUT** | **ENDES** | **NHANES** | **Health Survey for England** |
| **Survey years** | 2003, 2007, 2010 | 2006, 2012, 2018 | 2018-2020 | 1999-2000; 2001-02; 2003-04; 2005-06; 2007-08; 2009-10; 2011-12; 2013-14; 2015-16; 2017-18 | 1997-2019 |
| **Adults** | 17+ (2003); 15+ (2010, 2017) | 20+ | 16+ | 20+ | 16+ |
| **Survey design** | Nationally representative of urban and rural populations in 15 regions, drawn using probabilistic, geographically stratified, and multistage sampling | Nationally representative of people in community households, drawn using probabilistic, stratified, two-stage cluster sampling | Nationally representative (at the departmental level and by urban and rural area), drawn using two-stage, probabilistic, balanced, stratified sampling | Nationally representative of the US civilian non-institutionalized population, drawn using probabilistic, stratified, multistage, cluster sampling | Nationally representative of people living in private households, drawn using multistage stratified probability sampling |
| **Sampling of individuals (adults)** | One resident aged ≥15 years; oversampling of persons aged ≥65 years | One adult per household | One adult per household | A subsample of individuals selected based on sex, age, race and Hispanic origin, and income | All adults (maximum of 10) at selected address |
| **Data collection** | Face-to-face interviews followed by nurse visits at participants' home | Face-to-face interviews followed by nurse visits at participants' home | Face-to-face interviews followed by nurse visits at participants' home | Face-to-face interviews followed by visits to Mobile Examination Center (MEC) | Face-to-face interviews followed by nurse visits at participants' home |

| | | | | | |
|---|---|---|---|---|---|
| **Height** | Portable stadiometers to the nearest 0.1cm | Portable stadiometers with 0·1 cm precision. Two measurements, with a third taken if the difference was ≥1cm. Average of two measurements | Mobile, multipurpose wooden stadiometers or tall meter with a precision of 0.1cm. Average of three measurements | Wallmounted digital stadiometers, values recorded automatically. Head positioned in the Frankfort plane | Portable stadiometers with a sliding head plate, a base plate, and connecting rods marked with a measuring scale. Head positioned in the Frankfort plane |
| **Weight** | Barefoot and light clothing. Measured with a digital scale with an accuracy of 0.1kg | Fasting conditions and light clothing. Digital scales with 0.1kg precision. Two measurements, with a third taken if the difference was ≥400g. Average of the valid measurements | Digital scales with precision of 50 g | Participants wore the standard MEC examination gown. Digital scales to the nearest 0.1kg, values recorded automatically | Weight (in bare feet and minimal clothes). Digital scales to the nearest 0.1kg |
| **Waist circumference** | By an ergonomic circumference measuring tape. WC measured at the mid-axillary line at the midpoint between the costal margin and the iliac crest | By a flexible fibreglass anthropometric tape. WC measured at the midpoint between the highest part of the iliac crest and the lowest part of the ribs' margin of the median axial line. A | By a retractable metal tape. WC measured at the midpoint between the lower rib margin and the iliac crest. A resolution of 0.1cm | By a steel measuring tape. WC measured at the uppermost lateral border of the hip crest (ilium). A resolution of 0.1cm. | By a standard tape measure. WC measured at the midpoint between the lower rib and the upper margin of the iliac crest. Resolution to 0.1cm. |

| | | | | | |
|---|---|---|---|---|---|
| | | resolution of 0.1cm. Average of two measurements | | | Two measurements, with a third taken if they differed by more than 3cm. Average of the two closest valid measurements |

**ENS**: National Health Survey "*Encuesta Nacional de Salud*"; **HSE**: The Health Survey of England; **ENSANUT**: the National Health and Nutrition Survey "*Encuesta Nacional de Salud y Nutrición*"; **ENDES**: The Demographic and Family Health Survey *"Encuesta Demográfica y de Salud Familiar";* **NHANES**: The National Health and Nutrition Examination Survey

**Table S2A**. Sample characteristics (aged 25-64y) and anthropometric outcomes (height, weight, BMI and BMI status) by first and last survey periods (4-year cycle in US)

| | Chile | | Mexico | | Peru | | United States | | | | | | England | |
|---|---|---|---|---|---|---|---|---|---|---|---|---|---|---|
| | | | | | | | NH white | | NH black | | Mexican-American | | | |
| | 2003 | 2017 | 2006 | 2018 | 2018 | 2020 | 1999-2002 | 2015-18 | 1999-2002 | 2015-18 | 1999-2002 | 2015-18 | 1997 | 2019 |
| n | 2,159 | 3,414 | 25,006 | 12,083 | 23,992 | 15,615 | 2,501 | 2,054 | 1,169 | 1,628 | 1,405 | 1,159 | 5,499 | 4,316 |
| Age (years) | 44.8 | 45.6 | 41.1 | 43.3 | 39.8 | 40.6 | 44.8 | 44.8 | 44.2 | 45.9 | 43.6 | 45.1 | 43.2 | 45.1 |
| % Men | 45.9 | 35.3 | 39.7 | 43.4 | 43.5 | 44.6 | 51.3 | 50.0 | 47.7 | 46.0 | 50.3 | 47.4 | 47.3 | 44.2 |
| **Mean height (cm):** | | | | | | | | | | | | | | |
| *Men:* | | | | | | | | | | | | | | |
| Mean | 168.9 | 170.2[a] | 165.5 | 166.7[a] | 164.6 | 164.5 | 177.8 | 177.8 | 177.2 | 176.6 | 169.5 | 170.1 | 175.2 | 176.7[a,b] |
| SE | 0.28 | 0.31 | 0.14 | 0.16 | 0.13 | 0.16 | 0.20 | 0.26 | 0.22 | 0.31 | 0.30 | 0.56 | 0.15 | 0.21 |
| *Women:* | | | | | | | | | | | | | | |
| Mean | 155.5 | 156.5[a] | 152.5 | 154.0[a] | 152.0 | 152.2 | 163.8 | 163.8 | 163.7 | 163.1[a] | 157.5 | 156.7 | 161.8 | 162.7[a] |
| SE | 0.28 | 0.29 | 0.11 | 0.14 | 0.10 | 0.13 | 0.19 | 0.25 | 0.23 | 0.21 | 0.34 | 0.38 | 0.13 | 0.17 |
| **Mean weight (kg):** | | | | | | | | | | | | | | |
| *Men:* | | | | | | | | | | | | | | |
| Mean | 77.5 | 83.3[a] | 75.9 | 79.9[a] | 74.6 | 74.9 | 88.6 | 94.3[a] | 87.6 | 93.8[a] | 82.1 | 89.9[a] | 82.8 | 87.2[a] |
| SE | 0.53 | 0.64 | 0.25 | 0.35 | 0.29 | 0.33 | 0.56 | 0.92 | 0.73 | 0.96 | 0.44 | 0.97 | 0.29 | 0.47 |
| *Women:* | | | | | | | | | | | | | | |
| Mean | 67.8 | 73.5[a] | 67.9 | 71.0[a] | 65.5 | 66.2[a] | 74.7 | 79.3[a] | 83.5 | 87.1[a] | 72.8 | 78.4[a] | 69.1 | 74.0[a] |
| SE | 0.58 | 0.50 | 0.21 | 0.30 | 0.19 | 0.23 | 0.70 | 1.02 | 0.68 | 0.85 | 0.71 | 1.07 | 0.29 | 0.41 |
| **Mean BMI ($kg/m^2$)** | | | | | | | | | | | | | | |
| *Men:* | | | | | | | | | | | | | | |
| Mean | 27.1 | 28.7[a] | 27.7 | 28.7[a] | 27.5 | 27.6 | 28.0 | 29.8[a] | 27.8 | 30.0[a] | 28.5 | 31.0[a] | 27.0 | 27.9[a,b] |
| SE | 0.15 | 0.20 | 0.07 | 0.10 | 0.08 | 0.10 | 0.19 | 0.27 | 0.22 | 0.29 | 0.16 | 0.26 | 0.08 | 0.13 |
| *Women:* | | | | | | | | | | | | | | |
| Mean | 28.1 | 30.0[a] | 29.1 | 29.9[a] | 28.3 | 28.5 | 27.8 | 29.6[a] | 31.1 | 32.7[a] | 29.3 | 31.8[a] | 26.4 | 28.0[a] |
| SE | 0.26 | 0.20 | 0.08 | 0.12 | 0.07 | 0.09 | 0.28 | 0.38 | 0.25 | 0.31 | 0.34 | 0.40 | 0.11 | 0.15 |

| | Chile | | Mexico | | Peru | | United States | | | | | | England | |
|---|---|---|---|---|---|---|---|---|---|---|---|---|---|---|
| | | | | | | | NH white | | NH black | | Mexican-American | | | |
| | 2003 | 2017 | 2006 | 2018 | 2018 | 2020 | 1999-2002 | 2015-18 | 1999-2002 | 2015-18 | 1999-2002 | 2015-18 | 1997 | 2019 |
| **BMI status (%)** | | | | | | | | | | | | | | |
| *Men:* | | | | | | | | | | | | | | |
| Underweight | 0.3 | 0.6 | 0.9 | 0.5 | 0.2 | 0.3 | 1.0 | 0.7 | 1.8 | 1.1 | 0.2 | 0.0 | 0.3 | 0.5 |
| Normal weight | 27.4 | 17.1 | 27.2 | 21.7 | 30.6 | 28.0 | 29.4 | 23.8 | 33.2 | 22.2 | 21.7 | 10.8 | 33.1 | 28.3 |
| Overweight | 51.0 | 47.0 | 45.3 | 43.2 | 45.0 | 47.1 | 41.5 | 32.4 | 37.0 | 32.7 | 47.8 | 37.7 | 47.6 | 43.4 |
| Obesity I | 19.0 | 27.3 | 20.3 | 25.0 | 19.2 | 19.7 | 18.7 | 25.7 | 15.7 | 22.6 | 21.6 | 31.0 | 14.8 | 19.4 |
| Obesity II/III | 2.2 | 8.0 | 6.4 | 9.6 | 5.1 | 4.9 | 9.4 | 17.3 | 12.3 | 21.3 | 8.6 | 20.5 | 4.3 | 8.3 |
| *Overwt incl. obesity* | *72.2* | *82.3*[a] | *72.0* | *77.8*[a] | *69.3* | *71.7*[a] | *69.6* | *75.4*[a] | *65.0* | *76.6*[a] | *78.0* | *89.2*[a] | *66.7* | *71.1*[a,b] |
| *Obesity* | *21.2* | *35.3*[a] | *26.7* | *34.6*[a] | *24.3* | *24.6* | *28.1* | *43.0*[a,b] | *28.0* | *43.9*[a] | *30.2* | *51.5*[a] | *19.1* | *27.7*[a] |
| *Women:* | | | | | | | | | | | | | | |
| Underweight | 0.7 | 0.5 | 0.7 | 0.8 | 0.5 | 0.4 | 3.1 | 1.7 | 1.5 | 0.9 | 1.1 | 0.3 | 1.6 | 1.1 |
| Normal weight | 32.1 | 18.2 | 22.3 | 18.0 | 24.3 | 23.4 | 39.5 | 32.3 | 21.2 | 18.7 | 25.3 | 15.0 | 45.7 | 37.4 |
| Overweight | 35.2 | 37.9 | 37.8 | 37.3 | 42.8 | 42.4 | 25.8 | 26.2 | 27.7 | 22.4 | 34.2 | 32.1 | 32.4 | 30.0 |
| Obesity I | 22.1 | 25.2 | 25.7 | 26.9 | 24.1 | 23.7 | 16.0 | 18.2 | 23.2 | 21.9 | 24.0 | 23.1 | 12.8 | 18.3 |
| Obesity II/III | 9.9 | 18.2 | 13.5 | 17.0 | 8.3 | 10.1 | 15.7 | 21.5 | 26.4 | 36.1 | 15.4 | 29.6 | 7.6 | 13.2 |
| *Overwt incl. obesity* | *67.2* | *81.3*[a] | *77.0* | *81.2*[a] | *75.2* | *76.2* | *57.5* | *65.9*[a] | *77.3* | *80.4* | *73.6* | *84.8*[a] | *52.8* | *61.5*[a] |
| *Obesity* | *32.0* | *43.4*[a] | *39.2* | *43.9*[a] | *32.4* | *33.8* | *31.7* | *39.7*[a] | *49.6* | *58.0* | *39.4* | *52.7*[a] | *20.4* | *31.5*[a] |

*Abbreviations:* BMI: Body mass index; cm: centimetres; kg: kilograms; m: metres; n (unweighted sample size); SE: standard error. *Source (first and final year)*: Chile: Encuesta Nacional de Salud (ENS: 2003 and 2017); England: The Health Survey for England (1997 and 2019); Mexico: Encuesta Nacional de Salud y Nutrición (ENSANUT: 2006 and 2018); Peru: Encuesta Demográfica y de Salud Familiar (ENDES: 2018 and 2020); United States: The National Health and Nutrition Examination Survey (NHANES 1999-2002 and 2015-18). *Definitions:* Underweight (<18.5kg/m$^2$), normal weight (18.5 to 24.9kg/m$^2$), overweight (25.0-29.9kg/m$^2$), class I obesity (30.0 to 34.9kg/m$^2$), class II/III obesity (≥ 35.0kg/m$^2$), overweight including obesity (≥25.0kg/m$^2$), obesity (≥30.0kg/m$^2$). *Data:* Estimates for anthropometric variables are age-adjusted to the US year 2000 Census population using five-year age groups; estimates for mean age and % male are unweighted. Data are means (standard errors) or %.

[a]P<0.05 for difference in means separately by sex. [b]P<0.05: for sex difference in the means.

**Table S2B**. Sample characteristics (aged 25-64 years) and anthropometric outcomes (WC and abdominal obesity) by first and last survey periods (4-year cycle in US)

| | Chile | | Mexico | | Peru | | United States | | | | | | England | |
|---|---|---|---|---|---|---|---|---|---|---|---|---|---|---|
| | | | | | | | NH white | | NH black | | Mexican-American | | | |
| | **2003** | **2017** | **2006** | **2018** | **2018** | **2020** | **1999-2002** | **2015-18** | **1999-2002** | **2015-18** | **1999-2002** | **2015-18** | **1997** | **2019** |
| n | 2156 | 3395 | 24,705 | 11,972 | 23,691 | 15,416 | 2475 | 2008 | 1146 | 1559 | 1386 | 1119 | 4930 | 2716 |
| Age (years) | 44.8 | 45.6 | 41.1 | 43.2 | 39.8 | 40.6 | 44.9 | 44.8 | 44.2 | 46.0 | 43.6 | 45.1 | 43.3 | 45.7 |
| % Men | 45.8 | 35.4 | 39.6 | 43.5 | 43.6 | 44.9 | 51.5 | 50.2 | 47.6 | 46.5 | 50.1 | 47.5 | 47.4 | 43.0 |
| **Mean WC (cm):** | | | | | | | | | | | | | | |
| *Men:* | | | | | | | | | | | | | | |
| Mean | 92.9 | 97.1[a] | 95.3 | 98.4[a] | 94.9 | 94.9 | 99.9 | 103.9[a,b] | 95.7 | 101.1 | 98.9 | 104.3[a] | 95.4 | 97.4[a,b] |
| SE | 0.39 | 0.48 | 0.19 | 0.27 | 0.22 | 0.23 | 0.44 | 0.68 | 0.50 | 0.84 | 0.41 | 0.62 | 0.24 | 0.44 |
| *Women:* | | | | | | | | | | | | | | |
| Mean | 88.0 | 94.0[a,b] | 93.9 | 96.2[a] | 93.6 | 93.8 | 91.3 | 98.0[a] | 97.2 | 103.1 | 94.1 | 101.5[a] | 82.9 | 88.9[a] |
| SE | 0.57 | 0.47 | 0.20 | 0.29 | 0.17 | 0.21 | 0.71 | 0.87 | 0.66 | 0.77 | 0.83 | 0.91 | 0.27 | 0.45 |
| **Abdominal obesity (%):** | | | | | | | | | | | | | | |
| Men (>102cm) | 18.4 | 28.8[a] | 25.1 | 34.2[a] | 24.3 | 23.9 | 40.0 | 51.9[a] | 30.0 | 44.6[a,b] | 36.2 | 53.6[a] | 24.7 | 32.4[a,b] |
| Women (>88cm) | 49.6 | 67.2[a] | 66.6 | 74.0[a] | 68.5 | 67.9 | 53.6 | 66.8[a] | 70.1 | 77.3[a] | 64.2 | 81.5[a] | 28.5 | 46.9[a] |

*Abbreviations: cm: centimetres;* n (unweighted sample size); SE: standard error; WC: waist circumference. *Source (first and final year)*: Chile: Encuesta Nacional de Salud (ENS: 2003 and 2017); England: The Health Survey for England (1997 and 2019); Mexico: Encuesta Nacional de Salud y Nutrición (ENSANUT: 2006 and 2018); Peru: Encuesta Demográfica y de Salud Familiar (ENDES: 2018 and 2020); United States: The National Health and Nutrition Examination Survey (NHANES 1999-2002 and 2015-18).

*Estimates:* Estimates for anthropometric variables are age-adjusted to the US year 2000 Census population using five-year age groups; estimates for mean age and % male are unweighted. Data are means (standard errors) or %.

[a]P<0.05 for difference in means separately by sex. [b]P<0.05: for sex difference in the means.

**Supplemental Table 3A (Chile)** Predicted values at centiles of waist circumference (first and last) and difference (final minus first survey period) at cut points for overweight and obesity by sex

| WC (cm) | Chile | | | | | | |
|---|---|---|---|---|---|---|---|
| | Men | | | Women | | | Sex difference |
| | 2003, 2017 | Change β (95% CI) | P | 2003, 2017 | Change β (95% CI) | P | P |
| **BMI = 25kg/m$^2$** | | | | | | | |
| Mean | 86.3, 87.4 | 1.1 (0.3, 1.9) | 0.009 | 80.2, 82.1 | 1.8 (1.0, 2.7) | <0.001 | 0.145 |
| 5$^{th}$ | 80.0, 80.5 | 0.5 (-1.0, 2.0) | 0.503 | 73.5, 73.5 | -0.1 (-1.8, 1.6) | 0.943 | 0.621 |
| 25$^{th}$ | 83.4, 84.0 | 0.6 (-0.3, 1.5) | 0.198 | 76.9, 78.2 | 1.3 (0.4, 2.1) | 0.003 | 0.257 |
| 50$^{th}$ | 85.9, 86.7 | 0.9 (0.1, 1.7) | 0.032 | 79.3, 81.4 | 2.1 (1.1, 3.0) | <0.001 | 0.068 |
| 75$^{th}$ | 89.2, 90.1 | 0.9 (-0.3, 2.2) | 0.142 | 83.4, 86.1 | 2.7 (1.3, 4.1) | <0.001 | 0.060 |
| 95$^{th}$ | 92.9, 94.3 | 1.3 (-1.4, 4.0) | 0.335 | 89.1, 90.8 | 1.7 (0.7, 2.7) | 0.001 | 0.796 |
| **BMI = 30kg/m$^2$** | | | | | | | |
| Mean | 97.7, 98.3 | 0.6 (-0.2, 1.4) | 0.114 | 90.8, 93.0 | 2.1 (1.3, 3.0) | <0.001 | 0.009 |
| 5$^{th}$ | 91.6, 91.1 | -0.5 (-2.2, 1.3) | 0.611 | 83.3, 84.8 | 1.5 (-0.7, 3.7) | 0.173 | 0.169 |
| 25$^{th}$ | 94.5, 95.1 | 0.7 (-0.5, 1.8) | 0.250 | 87.2, 89.0 | 1.8 (1.0, 2.6) | <0.001 | 0.105 |
| 50$^{th}$ | 97.2, 98.0 | 0.7 (-0.3, 1.8) | 0.178 | 90.0, 92.0 | 2.1 (1.0, 3.1) | <0.001 | 0.082 |
| 75$^{th}$ | 100.3, 101.3 | 1.0 (-0.2, 2.2) | 0.092 | 94.4, 96.9 | 2.5 (1.1, 3.8) | <0.001 | 0.125 |
| 95$^{th}$ | 105.3, 105.7 | 0.4 (-1.9, 2.8) | 0.726 | 99.6, 102.5 | 2.9 (1.7, 4.1) | <0.001 | 0.068 |
| **BMI = 35kg/m$^2$** | | | | | | | |
| Mean | 109.0, 108.8 | -0.2 (-1.7, 1.3) | 0.789 | 100.3, 102.8 | 2.4 (1.5, 3.4) | <0.001 | 0.003 |
| 5$^{th}$ | 101.4, 100.3 | -1.2 (-3.0, 0.7) | 0.232 | 92.3, 94.3 | 1.9 (-2.0, 5.8) | 0.331 | 0.163 |
| 25$^{th}$ | 104.3, 105.1 | 0.9 (-1.3, 3.0) | 0.440 | 96.4, 98.4 | 2.0 (0.7, 3.2) | 0.002 | 0.373 |
| 50$^{th}$ | 107.6, 108.9 | 1.2 (-2.0, 4.4) | 0.450 | 99.7, 102.1 | 2.4 (0.9, 3.8) | 0.002 | 0.526 |
| 75$^{th}$ | 112.1, 112.1 | 0.0 (-3.3, 3.3) | 0.999 | 104.2, 106.9 | 2.6 (1.2, 4.1) | <0.001 | 0.152 |
| 95$^{th}$ | 119.3, 118.4 | -0.9 (-4.1, 2.3) | 0.578 | 109.0, 112.9 | 3.9 (2.3, 5.5) | <0.001 | 0.008 |

**Supplemental Table 3B (Mexico)**. Predicted values of waist circumference (first and last) and difference (final minus first survey period) across centiles of waist circumference relative to BMI at cut points for overweight and obesity by sex

| | Mexico | | | | | | |
|---|---|---|---|---|---|---|---|
| WC (cm) | Men | | | Women | | | Sex difference |
| | 2006, 2018 | Change β (95% CI) | P | 2006, 2018 | Change β (95% CI) | P | P |
| | | | | **BMI = 25kg/m$^2$** | | | |
| Mean | 87.9, 88.5 | 0.6 (0.2, 1.0) | 0.002 | 84.4, 85.1 | 0.7 (0.0, 1.4) | 0.053 | 0.819 |
| 5th | 79.6, 81.1 | 1.4 (0.7, 2.1) | <0.001 | 74.3, 76.3 | 1.9 (1.1, 2.8) | <0.001 | 0.362 |
| 25th | 84.1, 84.9 | 0.9 (0.4, 1.3) | <0.001 | 79.7, 80.9 | 1.2 (0.7, 1.6) | <0.001 | 0.376 |
| 50th | 87.3, 88.1 | 0.8 (0.3, 1.2) | 0.001 | 83.8, 84.2 | 0.4 (-0.1, 1.0) | 0.128 | 0.354 |
| 75th | 91.0, 91.2 | 0.2 (-0.2, 0.6) | 0.420 | 88.1, 88.0 | -0.1 (-0.6, 0.3) | 0.543 | 0.320 |
| 95th | 97.2, 96.2 | -1.0 (-1.9, -0.1) | 0.038 | 95.3, 94.1 | -1.2 (-2.5, 0.1) | 0.071 | 0.782 |
| | | | | **BMI = 30kg/m$^2$** | | | |
| Mean | 98.5, 99.9 | 1.4 (1.0, 1.8) | <0.001 | 94.1, 94.8 | 0.7 (0.2, 1.2) | 0.007 | 0.028 |
| 5th | 89.7, 91.7 | 2.0 (1.1, 2.8) | <0.001 | 83.4, 85.4 | 2.0 (1.3, 2.7) | <0.001 | 0.929 |
| 25th | 94.5, 96.1 | 1.5 (1.0, 2.1) | <0.001 | 89.1, 90.6 | 1.6 (1.1, 2.0) | <0.001 | 0.917 |
| 50th | 98.4, 99.8 | 1.4 (0.9, 1.9) | <0.001 | 93.7, 94.4 | 0.7 (0.2, 1.2) | 0.003 | 0.045 |
| 75th | 102.1, 103.2 | 1.2 (0.8. 1.6) | <0.001 | 98.3, 98.3 | 0.0 (-0.5, 0.4) | 0.984 | <0.001 |
| 95th | 107.4, 108.1 | 0.8 (-0.2, 1.7) | 0.137 | 105.4, 104.4 | -0.9 (-1.8, -0.1) | 0.033 | 0.012 |
| | | | | **BMI = 35kg/m$^2$** | | | |
| Mean | 109.3, 111.0 | 1.7 (0.9, 2.5) | <0.001 | 103.4, 104.3 | 0.9 (0.4, 1.5) | 0.002 | 0.107 |
| 5th | 98.2, 100.7 | 2.5 (0.2, 4.8) | 0.036 | 91.4, 93.7 | 2.4 (1.5, 3.3) | <0.001 | 0.927 |
| 25th | 104.6, 106.6 | 2.0 (1.0, 3.0) | <0.001 | 97.9, 99.8 | 2.0 (1.4, 2.6) | <0.001 | 0.954 |
| 50th | 109.5, 111.1 | 1.6 (0.7, 2.5) | 0.001 | 103.0, 104.1 | 1.2 (0.5, 1.8) | <0.001 | 0.438 |
| 75th | 113.5, 114.9 | 1.4 (0.8, 2.0) | <0.001 | 107.9, 108.5 | 0.6 (0.0, 1.1) | 0.043 | 0.057 |
| 95th | 120.1, 120.3 | 0.1 (-1.2, 1.4) | 0.850 | 116.2, 114.9 | -1.4 (-2.4, -0.4) | 0.007 | 0.073 |

**Supplemental Table S3C (Peru)**. Predicted values of waist circumference (first and last) and difference (final minus first survey period) across centiles of waist circumference relative to BMI at cut points for overweight and obesity by sex

| | Peru | | | | | | |
|---|---|---|---|---|---|---|---|
| WC (cm) | Men | | | Women | | | Sex difference |
| | 2018, 2020 | Change β (95% CI) | P | 2018, 2020 | Change β (95% CI) | P | P |
| | | | **BMI = 25kg/m$^2$** | | | | |
| Mean | 87.2, 87.0 | -0.3 (-0.5, 0.0) | 0.031 | 84.8, 84.4 | -0.4 (-0.7, -0.1) | 0.010 | 0.560 |
| 5$^{th}$ | 81.0, 80.8 | -0.2, (-0.7, 0.4) | 0.539 | 77.8, 77.4 | -0.4 (-1.1, 0.2) | 0.217 | 0.581 |
| 25$^{th}$ | 84.6, 84.4 | -0.2 (-0.5, -0.1) | 0.206 | 82.1, 81.8 | -0.3 (-0.6, 0.0) | 0.044 | 0.662 |
| 50$^{th}$ | 87.2, 86.7 | -0.5 (-0.8, -0.2) | 0.002 | 84.9, 84.3 | -0.5 (-0.9, -0.2) | 0.002 | 0.771 |
| 75$^{th}$ | 90.0, 89.4 | -0.5 (-0.9, -0.1) | 0.007 | 87.5, 87.2 | -0.3 (-0.6, 0.0) | 0.036 | 0.401 |
| 95$^{th}$ | 93.5, 93.4 | -0.1 (-0.7, 0.4) | 0.633 | 91.7, 91.2 | -0.4 (-1.0, 0.1) | 0.118 | 0.455 |
| | | | **BMI = 30kg/m$^2$** | | | | |
| Mean | 99.1, 98.7 | -0.4 (-0.7, -0.1) | 0.019 | 95.7, 95.1 | -0.5 (-0.8, -0.2) | <0.001 | 0.523 |
| 5$^{th}$ | 92.6, 92.3 | -0.2 (-1.0, 0.5) | 0.547 | 88.6, 87.7 | -0.9 (-1.5, -0.3) | 0.002 | 0.161 |
| 25$^{th}$ | 96.4, 96.1 | -0.3 (-0.7, 0.1) | 0.113 | 92.8, 92.3 | -0.5 (-0.9, -0.2) | 0.003 | 0.457 |
| 50$^{th}$ | 98.9, 98.5 | -0.5 (-0.9, -0.1) | 0.026 | 95.7, 94.9 | -0.8 (-1.2, -0.5) | <0.001 | 0.197 |
| 75$^{th}$ | 101.9, 101.4 | -0.5 (-1.1, 0.0) | 0.040 | 98.6, 98.0 | -0.5 (-0.8, -0.2) | 0.001 | 0.951 |
| 95$^{th}$ | 105.5, 105.1 | -0.4 (-0.8, 0.1) | 0.166 | 102.7, 102.5 | -0.2 (-0.7, 0.4) | 0.517 | 0.631 |
| | | | **BMI = 35kg/m$^2$** | | | | |
| Mean | 110.6, 109.6 | -0.9 (-1.5, -0.3) | 0.003 | 105.5, 104.9 | -0.5 (-0.9, -0.1) | 0.007 | 0.304 |
| 5$^{th}$ | 103.5, 103.3 | -0.2 (-2.1, 1.6) | 0.820 | 97.9, 97.0 | -0.8 (-1.5, -0.1) | 0.023 | 0.542 |
| 25$^{th}$ | 107.7, 106.7 | -1.0 (-1.6, -0.4) | 0.001 | 102.5, 101.9 | -0.6 (-1.1, -0.1) | 0.013 | 0.376 |
| 50$^{th}$ | 110.4, 109.4 | -1.0 (-1.7, -0.2) | 0.011 | 105.3, 104.6 | -0.6 (-1.3, 0.0) | 0.054 | 0.544 |
| 75$^{th}$ | 113.7, 112.8 | -0.9 (-2.1, 0.4) | 0.178 | 108.5, 108.0 | -0.5 (-0.9, -0.1) | 0.021 | 0.611 |
| 95$^{th}$ | 117.3, 116.4 | -0.9 (-1.9, 0.0) | 0.055 | 113.1, 112.7 | -0.4 (-1.2, 0.3) | 0.274 | 0.405 |

**Supplemental Table S3D (US non-Hispanic Whites)**. Predicted values of waist circumference (first and last) and difference (final minus first survey period) across centiles of waist circumference relative to BMI at cut points for overweight and obesity by sex

| | **US non-Hispanic Whites** | | | | | | |
|---|---|---|---|---|---|---|---|
| **WC (cm)** | **Men** | | | **Women** | | | **Sex difference** |
| | **1999-2002, 2015-2018** | **Change β (95% CI)** | **P** | **1999-2002, 2015-2018** | **Change β (95% CI)** | **P** | **P** |
| | | | | **BMI = 25kg/m$^2$** | | | |
| Mean | 90.6, 90.2 | -0.4 (-1.0, 0.3) | 0.261 | 83.7, 86.3 | 2.6 (1.8, 3.3) | <0.001 | <0.001 |
| 5th | 83.0, 83.1 | 0.1 (-0.9, 1.1) | 0.876 | 74.9, 78.3 | 3.5 (2.6, 4.4) | <0.001 | <0.001 |
| 25th | 87.1, 87.1 | 0.0 (-0.8, 0.8) | 0.998 | 80.0, 82.5 | 2.5 (1.7, 3.3) | <0.001 | <0.001 |
| 50th | 90.1, 90.1 | 0.0 (-0.6, 0.6) | 0.959 | 83.2, 85.9 | 2.8 (2.1, 3.4) | <0.001 | <0.001 |
| 75th | 93.6, 93.1 | -0.5 (-1.2, 0.2) | 0.155 | 87.4, 89.6 | 2.2 (1.3, 3.1) | <0.001 | <0.001 |
| 95th | 98.0, 97.3 | -0.8 (-2.1, 0.5) | 0.241 | 92.8, 95.3 | 2.5 (1.6, 3.3) | <0.001 | <0.001 |
| | | | | **BMI = 30kg/m$^2$** | | | |
| Mean | 103.2, 103.1 | -0.1 (-0.7, 0.4) | 0.622 | 94.8, 97.8 | 3.0 (2.1, 3.9) | <0.001 | <0.001 |
| 5th | 96.0, 95.5 | -0.6 (-1.5, 0.4) | 0.230 | 83.9, 88.7 | 4.8 (3.5, 6.2) | <0.001 | <0.001 |
| 25th | 99.7, 99.7 | 0.0 (-0.7, 0.7) | 0.940 | 90.2, 93.8 | 3.6 (2.6, 4.6) | <0.001 | <0.001 |
| 50th | 102.3, 102.7 | 0.3 (-0.3, 1.0) | 0.329 | 94.1, 97.4 | 3.3 (2.4, 4.1) | <0.001 | <0.001 |
| 75th | 106.6, 106.2 | -0.4 (-1.2, 0.3) | 0.249 | 99.0, 101.4 | 2.4 (1.4, 3.4) | <0.001 | <0.001 |
| 95th | 111.7, 111.1 | -0.5 (-1.6, 0.5) | 0.319 | 105.6, 107.7 | 2.1 (1, 3.2) | <0.001 | 0.001 |
| | | | | **BMI = 35kg/m$^2$** | | | |
| Mean | 115.2, 115.3 | 0.1 (-0.6, 0.9) | 0.737 | 105.0, 108.4 | 3.3 (2.2, 4.5) | <0.001 | <0.001 |
| 5th | 107.3, 107.2 | -0.2 (-2.4, 2.1) | 0.891 | 92.5, 97.4 | 4.9 (3.1, 6.7) | <0.001 | 0.001 |
| 25th | 111.9, 111.6 | -0.2 (-1.1, 0.7) | 0.639 | 99.7, 103.9 | 4.2 (2.8, 5.5) | <0.001 | <0.001 |
| 50th | 114.4, 114.7 | 0.3 (-0.5, 1.2) | 0.432 | 104.5, 108.2 | 3.7 (2.6, 4.8) | <0.001 | <0.001 |
| 75th | 119.0, 118.8 | -0.3 (-1.2, 0.6) | 0.522 | 109.9, 112.7 | 2.9 (1.6, 4.1) | <0.001 | <0.001 |
| 95th | 125.0, 123.9 | -1.1 (-2.3, 0.2) | 0.090 | 117.4, 119.5 | 2.2 (0.7, 3.7) | 0.005 | 0.001 |

**Supplemental Table S3E (US non-Hispanic Blacks)**. Predicted values of waist circumference (first and last) and difference (final minus first survey period) across centiles of waist circumference relative to BMI at cut points for overweight and obesity by sex

| WC (cm) | US non-Hispanic Blacks | | | | | | |
|---|---|---|---|---|---|---|---|
| | Men | | | Women | | | Sex difference |
| | 1999-2002, 2015-2018 | Change β (95% CI) | P | 1999-2002, 2015-2018 | Change β (95% CI) | P | P |
| **BMI = 25kg/m²** | | | | | | | |
| Mean | 85.9, 86.0 | 0.1 (-0.6, 0.8) | 0.733 | 82.7, 83.7 | 1.0 (0.0, 2.0) | 0.044 | 0.082 |
| 5th | 78.4, 78.4 | 0.0 (-1.8, 1.8) | 0.998 | 73.4, 75.4 | 2.0 (0.4, 3.6) | 0.012 | 0.102 |
| 25th | 82.6, 83.0 | 0.4 (-0.4, 1.2) | 0.334 | 79.1, 80.3 | 1.2 (0.2, 2.2) | 0.015 | 0.211 |
| 50th | 85.6, 85.5 | -0.2 (-0.9, 0.6) | 0.636 | 82.9, 83.6 | 0.7 (-0.5, 1.9) | 0.271 | 0.235 |
| 75th | 88.8, 89.1 | 0.4 (-0.7, 1.5) | 0.486 | 86.1, 87.0 | 0.9 (-0.2, 2.0) | 0.098 | 0.498 |
| 95th | 93.9, 93.5 | -0.4 (-1.6, 0.9) | 0.572 | 91.5, 92.2 | 0.7 (-1.1, 2.5) | 0.451 | 0.346 |
| **BMI = 30kg/m²** | | | | | | | |
| Mean | 98.6, 99.2 | 0.6 (-0.3, 1.4) | 0.177 | 93.5, 95.2 | 1.6 (0.6, 2.6) | 0.001 | 0.118 |
| 5th | 90.3, 90.1 | -0.2 (-1.8, 1.4) | 0.791 | 83.0, 85.5 | 2.4 (0.7, 4.1) | 0.005 | 0.028 |
| 25th | 94.9, 95.6 | 0.7 (-0.2, 1.7) | 0.130 | 89.7, 90.9 | 1.1 (0.2, 2.1) | 0.021 | 0.580 |
| 50th | 98.6, 98.9 | 0.3 (-0.6, 1.1) | 0.514 | 93.1, 94.9 | 1.8 (0.6, 3.1) | 0.004 | 0.044 |
| 75th | 101.7, 102.5 | 0.8 (-0.4, 2.1) | 0.199 | 97.3, 99.2 | 1.9 (0.6, 3.3) | 0.004 | 0.227 |
| 95th | 107.0, 107.1 | 0.1 (-1.5, 1.7) | 0.882 | 103.3, 105.2 | 1.8 (-0.6, 4.3) | 0.147 | 0.257 |
| **BMI = 35kg/m²** | | | | | | | |
| Mean | 110.9, 111.5 | 0.7 (-0.4, 1.8) | 0.223 | 103.4, 106.0 | 2.6 (1.4, 3.8) | <0.001 | 0.021 |
| 5th | 102.0, 101.1 | -1.0 (-2.6, 0.7) | 0.262 | 91.7, 94.8 | 3.1 (1.2, 5.0) | 0.001 | 0.002 |
| 25th | 106.6, 107.3 | 0.7 (-0.6, 1.9) | 0.281 | 99.2, 100.9 | 1.7 (0.6, 2.8) | 0.002 | 0.227 |
| 50th | 111.2, 111.7 | 0.5 (-0.5, 1.5) | 0.324 | 102.5, 105.8 | 3.3 (1.9, 4.8) | <0.001 | 0.002 |
| 75th | 114.2, 115.1 | 1.0 (-0.6, 2.5) | 0.210 | 107.6, 110.8 | 3.2 (1.7, 4.8) | <0.001 | 0.042 |
| 95th | 119.5, 120.2 | 0.7 (-1.9, 3.2) | 0.617 | 114.5, 117.2 | 2.8 (0.0, 5.5) | 0.046 | 0.271 |

**Supplemental Table S3F (US Mexican-Americans)**. Predicted values of waist circumference (first and last) and difference (final minus first survey period) across centiles of waist circumference relative to BMI at cut points for overweight and obesity by sex

| | **US Mexican-Americans** | | | | | | |
|---|---|---|---|---|---|---|---|
| **WC (cm)** | **Men** | | | **Women** | | | **Sex difference** |
| | **1999-2002, 2015-2018** | **Change β (95% CI)** | **P** | **1999-2002, 2015-2018** | **Change β (95% CI)** | **P** | **P** |
| | | | | **BMI = 25kg/m$^2$** | | | |
| Mean | 88.7, 88.7 | 0.0 (-0.7, 0.6) | 0.965 | 83.8, 85.9 | 2.0 (1.0, 3.0) | <0.001 | 0.002 |
| $5^{th}$ | 82.1, 81.4 | -0.7 (-2.4, 1.0) | 0.447 | 75.7, 78.5 | 2.8 (1.5, 4.2) | <0.001 | 0.002 |
| $25^{th}$ | 85.9, 86.0 | 0.1 (-0.9, 1.0) | 0.846 | 80.0, 82.3 | 2.3 (1.3, 3.3) | <0.001 | 0.002 |
| $50^{th}$ | 88.7, 88.6 | -0.1 (-1.2, 0.9) | 0.795 | 83.6, 85.9 | 2.3 (1.6, 3.1) | <0.001 | <0.001 |
| $75^{th}$ | 91.4, 91.5 | 0.1 (-0.9 ,1.0) | 0.904 | 87.1, 89.1 | 1.9 (0.8, 3.1) | 0.001 | 0.013 |
| $95^{th}$ | 94.6, 94.9 | 0.2 (-2.0, 2.5) | 0.838 | 91.3, 92.8 | 1.4 (-0.9, 3.7) | 0.225 | 0.463 |
| | | | | **BMI = 30kg/m$^2$** | | | |
| Mean | 101.2, 101.1 | -0.1 (-0.7, 0.5) | 0.779 | 94.5, 96.6 | 2.1 (0.8, 3.4) | 0.002 | 0.003 |
| $5^{th}$ | 94, 93.9 | -0.1 (-1.2, 1.0) | 0.859 | 85.2, 87.8 | 2.6 (1.3, 4.0) | <0.001 | 0.002 |
| $25^{th}$ | 98, 98.2 | 0.2 (-0.5, 0.9) | 0.547 | 90.7, 92.6 | 2 (0.7, 3.2) | 0.002 | 0.016 |
| $50^{th}$ | 101.1, 100.7 | -0.4 (-1.3, 0.6) | 0.425 | 94.3, 96.8 | 2.4 (1.5, 3.4) | <0.001 | <0.001 |
| $75^{th}$ | 103.8, 104.1 | 0.3 (-0.7, 1.4) | 0.546 | 98.7, 100.1 | 1.4 (0.6, 2.2) | 0.001 | 0.120 |
| $95^{th}$ | 108, 107.5 | -0.5 (-2.4, 1.3) | 0.567 | 102.6, 104.8 | 2.2 (-0.2, 4.7) | 0.077 | 0.078 |
| | | | | **BMI = 35kg/m$^2$** | | | |
| Mean | 113.1, 112.7 | -0.4 (-1.5, 0.7) | 0.456 | 104.5, 106.9 | 2.4 (0.9, 3.9) | 0.002 | 0.001 |
| $5^{th}$ | 105.2, 105.0 | -0.1 (-1.4, 1.1) | 0.837 | 93.8, 96.7 | 2.9 (1.4, 4.4) | <0.001 | 0.003 |
| $25^{th}$ | 109.7, 109.5 | -0.2 (-1.4, 0.9) | 0.714 | 100.4, 102.4 | 2.1 (0.5, 3.6) | 0.008 | 0.020 |
| $50^{th}$ | 112.9, 112.1 | -0.8 (-2.3, 0.6) | 0.273 | 104.4, 107.0 | 2.5 (1.3, 3.7) | <0.001 | 0.001 |
| $75^{th}$ | 115.7, 116.3 | 0.6 (-0.9, 2.1) | 0.414 | 109.4, 110.7 | 1.4 (0.4, 2.3) | 0.005 | 0.400 |
| $95^{th}$ | 121.3, 119.6 | -1.7 (-5.6, 2.1) | 0.381 | 113.1, 116.6 | 3.5 (0.6, 6.3) | 0.017 | 0.034 |

**Supplemental Table S3G (England)**. Predicted values of waist circumference (first and last) and difference (final minus first survey period) across centiles of waist circumference relative to BMI at cut points for overweight and obesity by sex

| | England | | | | | | |
|---|---|---|---|---|---|---|---|
| WC (cm) | Men | | | Women | | | Sex difference |
| | 1997, 2019 | Change β (95% CI) | P | 1997, 2019 | Change β (95% CI) | P | P |
| | | | | BMI = 25kg/m$^2$ | | | |
| Mean | 88.6, 89.1 | 0.5 (0.0, 1.0) | 0.064 | 77.9, 80.7 | 2.8 (2.3, 3.3) | <0.001 | <0.001 |
| 5$^{th}$ | 81.0, 81.5 | 0.5 (-0.5, 1.4) | 0.366 | 70.1, 72.6 | 2.5 (1.8, 3.1) | <0.001 | 0.001 |
| 25$^{th}$ | 85.5, 85.8 | 0.3 (-0.1, 0.7) | 0.146 | 74.3, 77.0 | 2.7 (2.3, 3.2) | <0.001 | <0.001 |
| 50$^{th}$ | 88.7, 88.9 | 0.2 (-0.4, 0.7) | 0.564 | 77.7, 80.7 | 3.0 (2.4, 3.5) | <0.001 | <0.001 |
| 75$^{th}$ | 91.7, 92.1 | 0.4 (-0.2, 1.0) | 0.169 | 81.2, 84.5 | 3.3 (2.8, 3.9) | <0.001 | <0.001 |
| 95$^{th}$ | 96.2, 97.8 | 1.7 (0.8, 2.5) | <0.001 | 86.8, 90.5 | 3.7 (3.0, 4.4) | <0.001 | 0.001 |
| | | | | BMI = 30kg/m$^2$ | | | |
| Mean | 100.2, 100.8 | 0.7 (0.1, 1.2) | 0.021 | 88.2, 91.7 | 3.5 (2.9, 4.2) | <0.001 | <0.001 |
| 5$^{th}$ | 92.4, 92.2 | -0.1 (-1.4, 1.1) | 0.852 | 78.8, 82.0 | 3.2 (2.5, 3.9) | <0.001 | 0.002 |
| 25$^{th}$ | 97.0, 97.4 | 0.4 (-0.3, 1.0) | 0.260 | 84.1, 87.4 | 3.3 (2.8, 3.8) | <0.001 | <0.001 |
| 50$^{th}$ | 100.3, 100.8 | 0.5 (-0.3, 1.2) | 0.219 | 88.0, 91.9 | 3.9 (3.0, 4.8) | <0.001 | <0.001 |
| 75$^{th}$ | 103.4, 104.1 | 0.6 (0.1, 1.1) | 0.017 | 92.2, 96.2 | 4.1 (3.4, 4.8) | <0.001 | <0.001 |
| 95$^{th}$ | 108.7, 110.4 | 1.7 (0.3, 3.1) | 0.016 | 98.5, 102.6 | 4.0 (3.0, 5.1) | <0.001 | 0.016 |
| | | | | BMI = 35kg/m$^2$ | | | |
| Mean | 111.8, 112.3 | 0.4 (-0.5, 1.4) | 0.382 | 98.1, 101.9 | 3.8 (3.0, 4.5) | <0.001 | <0.001 |
| 5$^{th}$ | 103.3, 102.9 | -0.4 (-1.8, 1.0) | 0.588 | 87.1, 90.3 | 3.3 (2.1, 4.4) | <0.001 | <0.001 |
| 25$^{th}$ | 108.3, 108.6 | 0.3 (-1.1, 1.7) | 0.690 | 93.5, 97.1 | 3.7 (3.0, 4.3) | <0.001 | <0.001 |
| 50$^{th}$ | 112.4, 112.6 | 0.2 (-1.2, 1.7) | 0.745 | 97.9, 102.3 | 4.4 (3.1, 5.7) | <0.001 | <0.001 |
| 75$^{th}$ | 115.8, 116.0 | 0.2 (-0.9, 1.3) | 0.719 | 102.7, 106.8 | 4.1 (3.2, 5.0) | <0.001 | <0.001 |
| 95$^{th}$ | 121.5, 122.6 | 1.1 (-0.8, 2.9) | 0.255 | 110.5, 113.7 | 3.2 (2.0, 4.5) | <0.001 | 0.023 |

*Source*: Chile: Encuesta Nacional de Salud (ENS: 2003, 2010, 2017); England: The Health Survey for England (1997 to 2019); Mexico: Encuesta Nacional de Salud y Nutrición (ENSANUT: 2006, 2012, 2018); Peru: Encuesta Demográfica y de Salud Familiar (ENDES: 2018, 2019 and 2020); US: The National Health and Nutrition Examination Survey (NHANES: 1999-2002; 2003-06; 2007-10; 2011-14; 2015-18).

Estimates shaded in grey are predicted values of mean waist circumference (first and last survey periods) and change over time (last minus first) at overweight and obesity cut-points. Calculated in linear regression analyses with survey year entered as a categorical variable (first year as referent). All models adjusted for age, age$^2$, BMI, and BMI$^2$ (age centred at 25y; BMI centred at 25kg/m$^2$) and included two three-way interaction terms (sex × year × BMI; sex × year × BMI$^2$) to allow the changes in mean WC relative to BMI to vary by sex.

Estimates not shaded are predicted values at centiles of waist circumference (first and last survey periods) and change over time (last minus first) at overweight and obesity cut-points. Calculated in quantile regression analyses with survey year entered as a categorical variable (first year as referent). All models adjusted for age, age$^2$, BMI, and BMI$^2$ (age centred at 25y; BMI centred at 25kg/m$^2$) and included two three-way interaction terms (sex × year × BMI; sex × year × BMI$^2$) to allow the centile-specific changes in WC relative to BMI to vary by sex.
